# VADEr: Vision Transformer-Inspired Framework for Polygenic Risk Reveals Underlying Genetic Heterogeneity in Prostate Cancer

**DOI:** 10.1101/2025.05.16.25327672

**Authors:** James V. Talwar, Adam Klie, Meghana S. Pagadala, Gil Pasternak, Brent Rose, Tyler M. Seibert, Melissa Gymrek, Hannah Carter

## Abstract

Polygenic risk scores (PRSs) serve as quantitative metrics of genetic liability for various conditions. Traditionally calculated as an effect size weighted genotype summation, this formulation assumes conditional feature independence and overlooks the potential for complex interactions among genetic variants. Transformers, a class of deep learning architectures known for capturing dependencies between features, have demonstrated remarkable predictive power across domains. In this work, we introduce VADEr, a Vision Transformer (ViT)-inspired architecture that combines techniques from both natural language processing and computer vision to capture properties exhibited by genetic data and model local and global interactions for genotype-to-phenotype prediction. Evaluating VADEr’s performance in predicting prostate cancer (PCa) risk, we found that across a range of metrics, including accuracy, average precision, and Matthews correlation coefficient, VADEr outperformed all benchmark methods, demonstrating its effectiveness in the context of complex disease risk prediction. To illuminate identified drivers of disease risk by VADEr, we formulated DARTH scores, an attention-based attribution metric, to capture the personalized contribution of each genomic region. These scores revealed distinct genetic heterogeneity captured by VADEr, with drivers of predicted risk identified in key PCa risk regions including the *HOXB13*, *TMPRSS2*, and *MSMB* loci. DARTH scores also revealed germline predispositions for particular PCa molecular subtypes, including an association between the *LMTK2* locus and the *SPOP* subtype, both implicated in the regulation of androgen receptor activity. Overall, by effectively capturing dependencies among genetic variants and providing interpretable insights, VADEr and DARTH scores offer a promising direction for advancing genotype-to-phenotype prediction, particularly in complex disease.

## Introduction

Decrypting genomic contributions to complex phenotypes remains a formidable challenge in precision medicine. Polygenic risk scores (PRSs), typically calculated as a genome-wide association study (GWAS) effect size weighted genotype summation, serve as tools for distilling genetic contributions into a metric of phenotype liability.^1^ However, such a formulation implicitly assumes an additive genetic architecture with risk variant independence,^2^ simplifying the complex dependencies and structured patterns that can exist in the genome.

Collapsing such complexities to a summation overlooks the rich dependency structures intrinsic to the genome; structures which closely parallel patterns in natural language processing (NLP) and computer vision (CV). Much like in language, where sequence dependencies and contextual relationships among words (or tokens) establish semantic meaning,^3^ genomic sequences display long-range dependencies between variants.^4–6^ Similarly, akin to the properties of spatial locality and compositionality found in CV problems, where local pixel patterns influence global image structure,^7^ genomic data demonstrates non-uniform statistical dependencies across loci.^8–10^ These hybrid characteristics, spanning both sequential and spatial dependencies, necessitate methods capable of capturing these properties, and highlight the potential of leveraging techniques from NLP and CV to enhance genomic modeling.

Transformers are a class of deep-learning architectures that have led to significant advancements in both of these domains.^11–15^ These models are modular in their structure, consisting of an alternating series of residual connection joined (multi-headed) attention layers and feedforward networks.^11,12^ Attention provides a mechanism for updating representations by considering pairwise similarity between any two elements in a sequence independent of distance, affording a powerful means of effectively capturing both short and long range dependencies. Awareness of the performance gains Transformer models can yield, coupled with recognition of the fundamental structural similarities the genome shares with language and images, has led to the widespread experimentation and application of Transformers to problems in genomics modeling.^5,16–18^

Efforts to directly link genetic variation to phenotypes using deep learning have spanned diverse datasets and traits.^19–23^ The outcomes of such studies, many of which employ simple and shallow architectures,^19–21^ have been mixed, showing success in non-complex phenotypes but limited utility for complex traits.^19,20,22,24^ Moreover, reported performance improvements are sometimes validated using data partitions rather than independent datasets, which may overstate model generalizability.^20,24–26^ Feature selection for dimensionality reduction, while essential for such modeling approaches, can similarly inflate performance when selection criteria are dataset-specific.^20^

Recent investigations utilizing Transformer architectures for genotype-to-phenotype prediction have reported promising results for several phenotypes, such as body mass index, systolic blood pressure, and lipid profiles.^24,26^ These successes, while encouraging, remain at the proof-of-concept stage, with validation performed on partitions of the same dataset rather than on independent external cohorts. Moreover, simplifying assumptions, such as grouping proximal variants to reach uniform dimensions and enable linear projection,^24^ likely yield suboptimal feature embeddings and reduce fidelity to the genomic structure.

Despite the potential of deep learning in genomic research, there are significant barriers to its clinical adoption for risk prediction. Interpretability is essential in clinical settings, and traditional PRS methods (including Bayesian shrinkage models)^9,27–30^ offer the advantage of inherent transparency.^10^ In contrast, while deep learning models offer theoretical performance advantages, their extensive parameter spaces and recursive non-linearities make it challenging to map feature contributions to model predictions. This proves prohibitive to identifying underlying biases that may compromise model generalizability, risking steep declines in performance when encountering out-of-distribution data.^31^ Moreover, extraction of novel biological insights identified by the model also remains a challenge. Ultimately, the absence of clear interpretability impedes trustworthiness, a critical factor in clinical contexts.^31,32^ Thus, interpretability should remain a key focus for next generation deep learning polygenic risk models.

Towards improving interpretability, inherently explainable deep learning architectures which hierarchically structure their network topology according to a defined biological prior have been proposed.^22,33^ Such architectures impose a relational inductive bias, similar to graph neural networks,^34^ and can yield performance gains, provided the imposed structure aligns with the underlying data distribution. However, as nodes and connections in these explainable architectures are defined from measured molecular interactions, which may be incomplete or restricted to certain biological contexts, such constraints can inadvertently limit model capacity. Designing these hierarchies also demands specialized expertise and custom architectures, reducing flexibility and scalability across biological domains.

Despite current challenges, Transformer-based architectures offer a compelling framework for advancing end-to-end genotype-to-phenotype prediction, particularly through their capacity to model context-dependent variant dependencies in conditions with heterogeneous risk profiles. Here we introduce the **V**ision **A**dapted **D**isease **E**lucidating T**r**ansformer (VADEr), an architecture inspired by Vision Transformers (ViTs) that combines techniques from NLP embeddings and ViT patch partitioning to generate chromosomally consistent, region-specific representations. As proof-of-concept, we assessed its performance in prostate cancer (PCa) risk prediction, ensuring independent test set evaluation. Across a range of metrics VADEr outperformed all benchmarks, including statistical genomics approaches with access to a much larger effective pool of information, demonstrating its potential for complex disease risk prediction. For interpretability, we formulated **D**irected **A**ttention **R**elevance from **T**ransformer **H**euristics (DARTH) scores, an attention-based attribution metric that captures personalized regional contributions driving predicted disease risk by VADEr. Analyses of these scores demonstrated VADEr’s capability to capture genetic heterogeneity and identify key patterns of attributed PCa risk, including the well-established *HOXB13* locus and the *TMPRSS2* region.^35–38^ DARTH scores also highlighted the potential of VADEr to uncover germline risk patterns associated with subtype-specific disease, exemplified by an identified association between the *LMTK2* locus and the *SPOP* PCa subtype in The Cancer Genome Atlas (TCGA) PCa individuals.^39^ Taken together, these results underscore the potential of the DARTH-VADEr framework for accurate and interpretable genotype-to-phenotype modeling and supports a broader exploration of this approach across genotype-to-phenotype prediction challenges.

## Methods

### Datasets

Genotypes and case/control status were downloaded for 91,644 male individuals (n_cases_ = 54,283; n_controls_ = 37,361) from the ELucidating Loci Involved in Prostate cancer SuscEptibility (ELLIPSE) Consortium via dbGaP (study accession: phs001120.v1.p1). Training and validation sets corresponded to a random 80/20 split of ELLIPSE, ensuring consistency in the frequencies of case-control status, ancestry, and family history (where available).

Ancestry was determined from the top 10 principal components computed with FlashPCA2^40^ on 1000 Genomes and ELLIPSE individuals. These top principal components were used as features for an ancestral k-means clustering, where fitting was performed on 1000 Genomes individuals (with each cluster corresponding to a specific ancestry). ELLIPSE ancestry was then assigned according to the nearest cluster. This procedure yielded a 99.2% agreement with self-reported ancestries in the ELLIPSE cohort. Overall, ELLIPSE exhibited a large overrepresentation of European ancestry individuals (n_European_ = 82,594) corresponding to 90.1% of the dataset, despite ancestry being a documented risk factor for PCa.^41,42^

To evaluate model generalizability, we constructed a balanced independent test set (n = 15,235; n_cases_ = 7,591; n_controls_ = 7,644) from a subsampling of the UK Biobank (UKBB). Specifically, test set cases were defined as all male individuals of European ancestry in the UKBB with an ICD code specific to PCa. Test set controls were randomly selected from all remaining male UKBB individuals of European ancestry without a cancer ICD code in an age-matched manner. Details of UKBB ancestry assignment can be found in the works of Richard *et al.*^43^ and Pagadala *et al.*^44^

To assess VADEr’s ability to capture germline risk associated with molecular subtypes, we utilized genotypes for European ancestry PCa individuals (n = 412) from TCGA, as downloaded and processed in previous work.^44^ Molecular subtype annotations for a subset of these individuals (n = 275) were acquired directly from the TCGA Research Network report^39^ on primary PCa. TCGA ancestry assignment procedure can be found in Pagadala *et al.*^44^

ELLIPSE, TCGA, and the UKBB differ in their genotyping and imputation methodologies. ELLIPSE was genotyped using the OncoArray microarray,^45^ while TCGA and the UKBB were genotyped using the Affymetrix SNP 6.0 Array^46^ and UK Biobank Axiom Array,^47^ respectively. For ELLIPSE, we imputed genotypes with Minimac4 using the 1000 Genomes Phase 3 Version 5 reference panel, by means of the Michigan Imputation Server.^48^ Quality control prior to ELLIPSE imputation is documented by Pagadala *et al.*^42^ TCGA genotypes were also imputed with Minimac4 via the TOPMed Imputation Server,^48^ but used the TOPMed-r3 reference panel. After imputation, TCGA variants were mapped from GRCh38 to GRCh37 coordinates using LiftOver.^49^ Full details of UKBB imputation can be found in the original UKBB report by Bycroft *et al*.^47^ Following imputation, ELLIPSE, TCGA, and the UKBB contained genotypes for 48,899,508, 444,576,000, and 97,013,422 variants respectively. All datasets and variants used in this study employed GRCh37 as the reference genome.

### Formulating Feature Sets

Given the high-dimensionality of the feature space, strong correlation that exists between features (i.e., variants) due to linkage disequilibrium (LD), and noise introduced by genotyping and imputation error, using all shared variants across datasets poses several problems for any PRS methodology. Classical additive PRSs, commonly known as clumping + thresholding (C+T) approaches, narrow the feature space by identifying and selecting the most significant GWAS variants at a locus and omitting all less significant correlates.^10^ Thresholding is then applied across a range of GWAS p-values, further reducing the feature set and creating a candidate set of scores, with the final PRS selected based on its best performance in an independent dataset.

Rather than omitting features, a number of PRS methods instead leverage LD-structure by reweighting single nucleotide polymorphism (SNP) effects according to Bayesian shrinkage techniques^9,27–30^ frequently yielding improved performances relative to C+T approaches. Though theoretically all variants could be used with these approaches, it is recommended to constrain the feature space to HapMap3 variants. Both classical and Bayesian PRSs are fundamentally dependent on GWAS summary statistics.

For machine learning (ML) based approaches, feature selection through an analogous C+T technique offers a reasonable tradeoff between reducing dimensionality and maintaining predictive power, making it a practical strategy for handling complex genomic data. To identify significant variants and their effect sizes, not only for p-value-based feature selection in VADEr and baseline ML models, but also to support the generation of both statistical genomics and classical PRSs, we downloaded summary statistics from dbGaP (study accession: phs001120.v2.p1) for a large-scale PCa multi-ancestry GWAS meta-analysis by Conti *et al.*^50^ It should be emphasized that ELLIPSE was a subset of this GWAS, but, perhaps more importantly, the UKBB was not, preventing any test set leakage during the feature selection phase.

Using LD structure from the 1000 Genomes dataset to determine SNP correlation, we performed clumping of summary statistics from *Conti et al.* at each associated locus using a radius of 125kb and an r^2^ of 0.8. P-value thresholds of 5e-8, 5e-7, 5e-6, 5e-5 and 5e-4 were subsequently applied to formulate candidate feature sets of 4,062, 5,231, 7,024, 10,389, and 12,017 variants, respectively. All variants employed for training, validating, and testing VADEr and baseline models passed classical quality control metrics including a minor allele frequency (MAF) exceeding 1% and an imputation quality score exceeding 0.8.^10^

Features for all models were encoded as alternate allele counts. Moreover, for VADEr and ML baseline models, these counts were standardized according to the mean and standard deviation of the training set distribution to improve training convergence. Finally, to ensure maximal feature recovery and orientation consistency across genomic datasets and summary statistics, all datasets were harmonized with GRIEVOUS.^51^

### VADEr Architecture

Similar to Vision Transformers (ViTs),^12^ we partition the genome into non-overlapping patches of length W, ensuring each patch is chromosomally consistent (i.e., all positions within a patch are located on the same chromosome, and do not overflow into another). All variants in a feature set are then mapped onto these patches, with strongly correlated features, as identified through the clumping procedure (Methods: *Formulating Feature Sets)*, removed. Across p-value thresholds (5e-8, 5e-7, 5e-6, 5e-5, 5e-4), this procedure yielded 331, 436, 603, 1,008, and 1,204 genomic patches, respectively. For the largest feature set (i.e., p = 5e-4), patch density ranged from 1 to 258 SNPs (*μ* = 9.98; *σ* = 19.6; median = 3).

Given the variable number of variants across patches, mapping each genomic patch to a constant latent vector size D with a single trainable linear projection, as is classically done, is infeasible. Instead, VADEr generates patch embeddings using independent trainable transformations, assigning a distinct linear layer to each patch. Although this increases the number of trainable parameters relative to a single shared projection, it enables richer customized regional embeddings (by effectively tokenizing each patch), which is particularly advantageous given the datatype (SNPs) and feature encodings (counts). Moreover, to account for non-linear interactions that may occur between variants within a patch, we applied GELU activation to each patch’s D-dimensional representation after projection.

With all genomic patches having been mapped to a D dimensional representation, we prepend a learnable classification token (CLS) and add learnable positional embeddings. We also append r learnable registers to the sequence of embedded patches. Registers were included to both improve performance and interpretability, as their inclusion has been shown to prevent artifacts attributable to token recycling from appearing in attention maps in ViT networks.^52^ This sequence of embedded vectors is then passed as input to a Transformer encoder.

The Transformer encoder^11,12^ consists of L layers, alternating between multi-headed self-attention (MSA) and a multi-layer perceptron (MLP). Within each layer, layer normalization (LN) is applied prior to MSA and after the residual connection (prior to the MLP) within each block.^12,53,54^ VADEr utilizes SwiGLU as its MLP nonlinear activation function.^55^ VADEr also employs a learnable temperature within each MSA head, to allow for a sharpening of the attention score distribution, which has been shown to be helpful when training ViTs from scratch on small-size datasets.^56^

Following the transformation of all patches through the entirety of the Transformer encoder, VADEr distills the underlying output to a single, informative individual representation using CLS pooling. This representation is then fed into a linear classification head for disease prediction. A detailed overview of the VADEr network architecture can be found in **Figure 1**.

**Figure 1:**
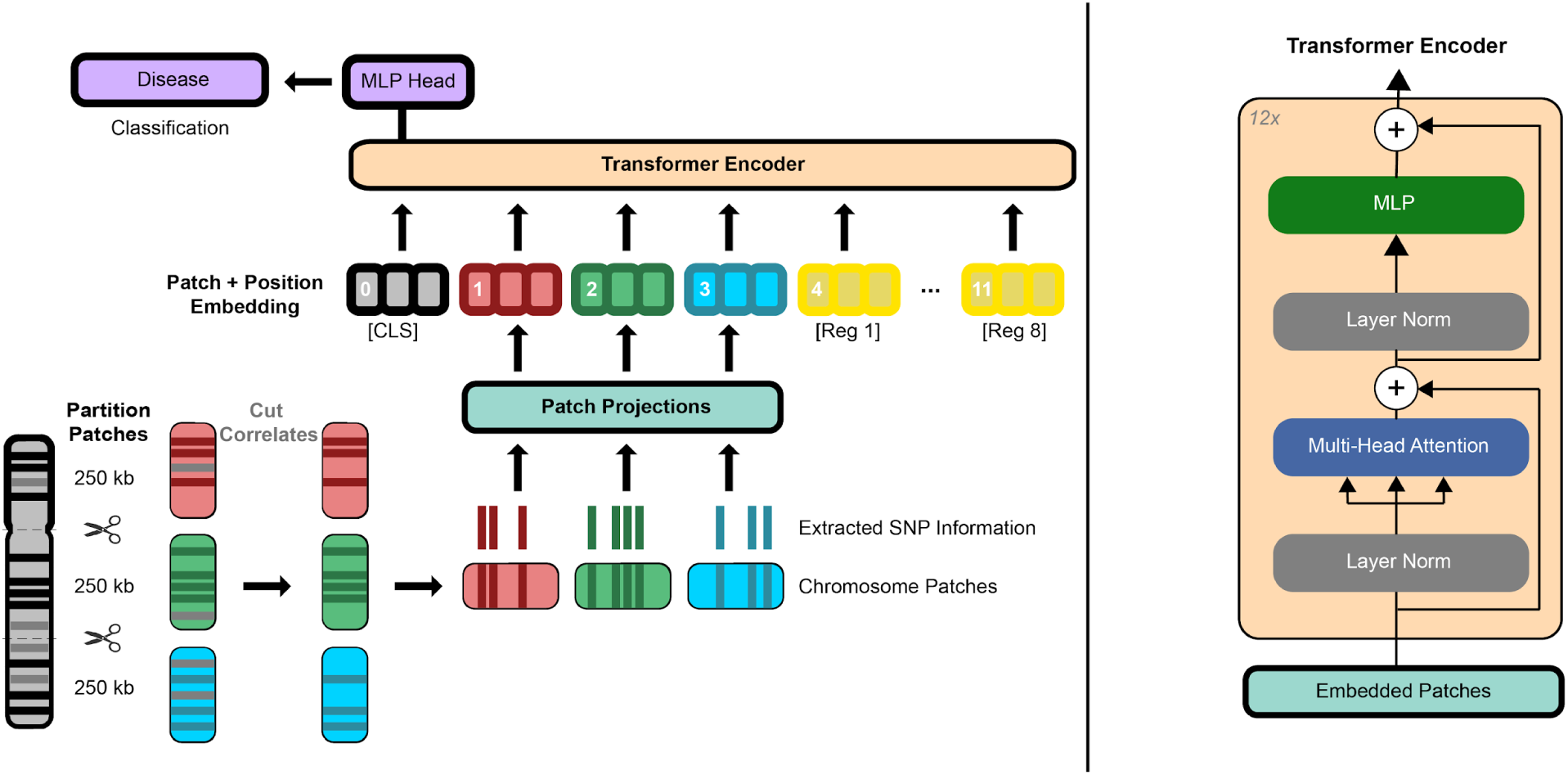
VADEr model overview. Each chromosome is partitioned into non-overlapping patches of 250kb. For each patch, variants in the candidate feature set (after p-value thresholding and clumping) are extracted and passed to a patch specific projection. Positional embeddings are subsequently added to the output of these projections. A learnable classification token (CLS) and registers are then prepended and appended to the representation, respectively. The resulting genomic sequence representation is then passed through a Transformer encoder, CLS pooled, and passed to a classification head for disease prediction.

Hyperparameters were selected through empirical evaluation of validation set performance, a detailed summary of which can be found in **Table 1**.

**Table 1:** Summary of VADEr model hyperparameters.

| Hyperparameter | Value |
| --- | --- |
| Patch Window Length (W) | 250kb |
| Embedding Dimension (D) | 768 |
| Number of Transformer Layers (L) | 12 |
| Number of Attention Heads | 12 |
| Number of Registers (r) | 8 |
| Patch Projection Dropout | 0.2 |
| Transformer Dropout | 0.2 |
| Patch Projection Activation | GELU |
| MLP Activation | SwiGLU |
| MLP Dimension | 2,048 |
| Query, Key, Value Dimension ($d_h$) | 64 |
| Learnable Temperature Initialization ($\tau$) | 8 |

### Training VADEr

VADEr models across all feature sets were trained using the same procedure, employing binary cross-entropy (BCE) as the loss function and AdamW^57^ as the optimizer, with β_1_ = 0.9, β_2_ = 0.99, ε = 10^−8^, and a weight decay of 0.1. The learning rate was linearly warmed up to 8×10^−5^ over the first 95 steps (approximately 5 epochs), before being decayed according to a cosine schedule for the remainder of the 115 total epochs. To improve training stability, we applied gradient clipping at a global L2 norm of 10.

Model checkpointing occurred after every epoch, with metrics computed and stored for evaluation. For each feature set, the model that achieved the highest area under the receiver operating characteristic curve (AUC) on the validation set was selected as the best model.

All VADEr models were implemented in PyTorch and trained with an effective batch size of 4,096. To achieve this, we utilized a combination of distributed training and gradient accumulation. Distributed training was conducted across 4 A30 GPUs using PyTorch’s Distributed Data Parallel (DDP), with a local batch size set to the maximum even number of samples that could fit into GPU memory.

### Baselines

To contextualize VADEr’s performance, we optimized a number of baseline models spanning classical (i.e., C+T), Bayesian, and multiple ML approaches. We also implemented the 269 variant PCa PRS reported by Conti *et al.*,^50^ the large-scale PCa GWAS meta-analysis from which we obtained our summary statistics.

Classical PRS were constructed with PRSice-2.^58^ As with VADEr, p-value thresholding was conducted across 5e-8, 5e-7, 5e-6, 5e-5 and 5e-4, and the 1000 Genomes was used as the LD reference. The optimal classical PRS was identified as the threshold which maximized the validation set performance (AUC).

LDpred2^30^ was used as the Bayesian benchmark. As recommended by the authors, we constrained features to HapMap3+ variants.^59^ Employing the 1000 Genomes as the reference panel, we observed a marked divergence between validation (AUC = 0.83916) and test set performance (AUC = 0.66622), and a notable test set underperformance relative to C+T (**Table 2**). While the former may reflect that the validation set was used in the generation of the summary statistics by Conti *et al.*^50^, the latter suggests poor effect size reestimation, which is dependent upon the LD reference. Given the dependence of reestimation from LD information, we decided to use the entirety of ELLIPSE as the reference panel for LDpred2. Given that ELLIPSE made up 39.1% of the original study, we hypothesized that it would serve as a more suitable and representative LD reference, though this came at the cost of independently assessing hyperparameter performance on the validation set. Fortunately, LDpred2-auto is able to directly infer hyperparameter values, and thus this was the approach used and reported here. We note that though LDpred2 does provide LD matrices for use in cases such as this, these matrices are computed from European individuals of the UKBB, which if used here, would result in test set leakage.

**Table 2:** Held out test set performances of VADEr and baseline models across multiple performance metrics. P ≤ corresponds to the p-value threshold for which validation set performance was maximized. This was used to select the model for final evaluation on the test set. The best performing model for each metric is **bolded**, while the second-best model is <u>underlined</u>. ^➕^ denotes method utilized reported summary statistics’ effect sizes, which were inferred with 3.2x the amount of data as our training set. ^‡^ LDpred2 hyperparameter which corresponds to the proportion of causal variants.

| *Method* | *P* ≤ | # Variants | *AUC* | *Accuracy* | *AP* | *Precision* | *Recall* | *F1 Score* | *MCC* |
| --- | --- | --- | --- | --- | --- | --- | --- | --- | --- |
| C+T | 5e-5 | 730 | 0.68427 | 0.63853 | 0.67102 | 0.63875 | 0.63193 | 0.63532 | 0.27704 |
| LDpred2 | 0.0056<sup>‡</sup> | 965,472 | 0.71266 | 0.65711 | 0.69870 | <u>0.65880</u> | 0.64682 | 0.65275 | 0.31421 |
| Logistic Regression | 5e-4 | 12,017 | **0.72843** | 0.64687 | 0.71322 | 0.60590 | 0.83322 | **0.70161** | 0.31761 |
| SVM | 5e-4 | 12,017 | 0.72176 | 0.65369 | 0.70652 | 0.61878 | 0.79436 | 0.69566 | 0.32114 |
| Random Forest | 5e-5 | 10,389 | 0.67829 | 0.50594 | 0.67771 | 0.50212 | **0.99657** | 0.66778 | 0.07296 |
| XGB | 5e-4 | 12,017 | 0.72635 | 0.64102 | <u>0.71323</u> | 0.59944 | <u>0.84258</u> | <u>0.70051</u> | 0.30937 |
| FC-FFN | 5e-5 | 10,389 | 0.72525 | 0.65172 | 0.70903 | 0.61521 | 0.80371 | 0.69694 | 0.31940 |
| VADeR | 5e-4 | 12,017 | <u>0.72665</u> | **0.66400** | **0.71352** | 0.64931 | 0.70808 | 0.67742 | **0.32954** |
| *Conti et al.*<sup>‡50</sup> | N/A | 269 | 0.70611 | 0.64693 | 0.69208 | 0.64747 | 0.63970 | 0.64356 | 0.29385 |
| C + T<sup>+</sup> | 5e-4 | 2,894 | 0.68677 | 0.63328 | 0.67246 | 0.63060 | 0.63733 | 0.63395 | 0.26659 |
| LDpred2<sup>+</sup> | 0.032<sup>‡</sup> | 965,472 | 0.71949 | <u>0.66157</u> | 0.70528 | **0.65976** | 0.66236 | 0.66106 | <u>0.32314</u> |

It is important to highlight that PRSs generated with either C+T or LDpred2 make use of the effect sizes provided by summary statistics, while VADEr and the ML baselines (described subsequently), must learn a mapping between features and labels from the training set alone. As the training set size (n = 73,315) and the number of individuals used to generate the summary statistics in the Conti *et al.* study (n = 234,253) differ,^50^ comparison of models is not entirely “fair”, as C+T and LDpred2 have access to a larger pool of information. To obtain more equivalent models, we ran both PRSice-2 and LDpred2 again on summary statistics generated from our training set alone. These followed the same procedures as above, with the exception that for LDpred2 we used only the training set genotypes (as opposed to all of ELLIPSE) as the LD reference panel. This approach ensures a fair cross-method comparison of different approaches. For comprehensiveness though, we report results for C+T and LDpred2 for both the Conti *et al.* summary statistics and the training set summary statistics, with the former being denoted by a ^➕^ symbol.

ML baselines consisted of logistic regressions, support vector machines, random forests, XGBoosts, and fully-connected feedforward neural networks (FC-FFN). Hyperparameters for each approach (e.g., regularization, model depth) were optimized to maximize the validation set performance (AUC) using a Tree-structured Parzen Estimator (TPE) sampler with Optuna.^60^ Specifically, 100 trials were conducted across each p-value threshold feature set, resulting in 500 total trained models per baseline. The final model for each baseline ML approach was the one that resulted in the maximum validation AUC. Logistic regression, support vector machine, and random forest models were implemented with scikit-learn. XGBoost models were implemented with CUDA acceleration (on an A30 GPU) via the xgboost Python package. FC-FFN models were implemented in PyTorch, with each model utilizing a RTX 5000 GPU for training and inference.

Finally, all PRSs without an explicit boundary for classification (i.e., C+T, LDpred2, and the Conti *et al.* PRS), were transformed to a [0, 1] prediction probability via a logistic regression fit on top of the original method test set scores.

### DARTH Patch Attribution

Self-attention, or the assignment of pairwise attention between all token pairs, is the defining building block of Transformer networks. Directly utilizing attention values as a metric of relevance (often from the last layer in the model) for Transformer explainability is a common practice as it allows for an easy and intuitive identification of attention patterns. Though convenient, this approach fails to globally capture token contributions, omitting not only intermediate attention scores, but also the contributions of other model components.^61^

Methods such as attention rollout and attention flow^62^ attempt to improve attention based Transformer explainability by accounting for the intrinsic mixing of information across layers. However, each of these approaches suffers from limitations. Rollout, which linearly combines attentions along paths in the pairwise attention graph, fails to distinguish between positive and negative contributions to the decision, which leads to artifacts in the attention attribution.^61^ Attention flow, which conceptualizes the attention graph as a flow network and assigns attributions according to a max-flow problem formulation, can yield improved attributions relative to rollout in specific cases, but is too slow to support large-scale evaluations.

It is also worth emphasizing that each of these methods (i.e., attention maps, rollout, and flow along with many other non-attention based attribution approaches) are class-agnostic. Instead, we implemented an extension of Beyond Attention, a Transformer interpretability method proposed by Chefer *et al.*^61,63^ This approach provides a class-specific comprehensive capturing of information propagation through the network, and has been shown to outperform other interpretability approaches across multiple modalities.^63^

Patch relevance can be conceptualized as a self-contained entity prior to passing through VADEr’s Transformer encoder, as at this point each patch remains a local representation, independent of the information contained in any other patch. Given this we can initialize a relevance map R_0_ according to this self-containment principle as an identity matrix:

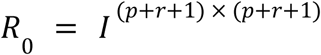

where p is the number of genomic patches in a given feature set. As patches pass through the Transformer, attention heads in each layer contextualize patch representations. Thus in any given layer, the attention maps can be used to update the relevance map, but must necessarily account for the varying importance and relevance of each attention head.^64^ To generate a layer-specific attention map 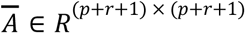, one can perform gradient-weighted averaging across heads:

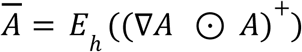

where

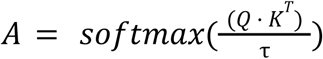

is the attention map of a given attention head, ⊙ is the Hadamard product, 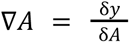 with y representing the model output, and *E_h_* is the mean across attention heads. Negative contributions are removed prior to averaging to capture positive relevance in accordance with the original method.^61,63^

The circuitry of relevance within a Transformer layer is also branching, with residual connections subsequent to each MSA operation. Thus appropriately updating R_i_ at each layer requires accounting for this facet of the architecture and leads to the formulation of the following relevancy update rule:

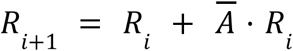

With this rule in place, we can compute personalized per patch **D**irected **A**ttention **R**elevance from **T**ransformer **H**euristics (DARTH) scores through the extraction of the CLS token row in R_L_.

### Perturbing Patches

For each DARTH score signature cluster identified (Results: *Linking VADEr Attributable Risk to Loci With DARTH Scores*) a matched size complement from a random sampling of non-cluster individuals was constructed. With both a cluster and its assigned complement established, patch-level genotypes were then shuffled independently both within a set and between sets. Using the dominating cluster 1 *HOXB13* patch and cluster 1 and its complement as an example, we would first shuffle only the genotypes captured by this patch among individuals in each set (intra-cluster). Next we would randomly shuffle this *HOXB13* patch genotype between individuals in each cluster (inter-cluster), ensuring all individuals in cluster 1 and its complement received a unique *HOXB13* patch genotype from outside their defined set. With both intra- and inter-cluster shuffling completed, a patch’s contribution to model prediction could be evaluated and contextualized by computing the difference in VADEr prediction probability for each patch-shuffled genotype relative to their original genotype.

### Statistical Analyses

For patch perturbation analyses, two-sided Mann-Whitney U tests were used to compare if prediction score changes came from the same distribution. Fisher’s exact tests were used to assess whether the number of VADEr-predicted cases differed significantly across attention-derived clusters. Statistical tests were implemented with the scipy.stats Python package. Wherever multiple hypotheses were tested, p-values were corrected by the Benjamini-Hochberg procedure. Permutation testing, as employed for DARTH score-molecular subtype investigations, utilized 1,000 unique random label permutations, preserving the original class balance of the defined subtype. In all instances where exact p-values could not be computed due to numerical underflow, results are reported as p ≪ 0.001.

## Results

### Evaluating VADEr’s Prognostic Potential

Transformers and their derivatives have emerged as the state-of-the-art across NLP benchmarks^11^ and have seen widespread adoption in both NLP and CV tasks. Their success is largely attributable to their self-attention mechanism, which allows for capturing complex, contextual relationships within data to effectively model both local and global patterns. Given the genome shares many intrinsic properties with data of these domains (e.g., sequential structures, long-range dependencies, non-uniform statistical relationships) and the demonstrated promise of Transformer-based approaches in genomic applications,^5,16–18^ we explored their use for complex disease risk prediction.

A significant barrier exists though between conceptualization and effective implementation. The genome’s prodigious size proves prohibitive to tractably computationally model the confluence of global genetic liability to a phenotype. Thus, rather than employing the whole genome as the feature space, it is instead common practice to focus on genomic differences, particularly SNPs, which are variations at specific loci. Though focusing on SNPs markedly reduces the data size, it introduces regions of varying density, complicating the application of classic modeling techniques that assume uniform data dimensions.

Recognizing these challenges and the need for specialized solutions, we developed VADEr, a ViT-inspired^12^ model architecture for complex disease risk prediction (**Figure 1**). VADEr conceptualizes the genome as an image which can be partitioned into regions, or patches, of static length, but of differing SNP density. Unlike classic ViTs which project all image patches (of equal pixel density) through a single trainable linear projection, VADEr projects each genomic patch through its own single layer non-linear projection to generate a regional representation of consistent dimensionality (Methods: *VADEr Architecture*). These patch representations are then passed through a Transformer encoder, CLS pooled, and passed to a final classification head to generate a disease prediction.

We evaluated VADEr’s potential for complex disease prediction in prostate cancer (PCa), a highly heritable complex condition for which a large non-private case-control genotype dataset exists. From ELLIPSE, we constructed training and validation sets according to a random 80/20 split (Methods: *Datasets*). To reduce the feature space for model training, we employed a clumping and thresholding based approach employing linkage structure from the 1000 Genomes^65^ and PCa GWAS p-values from Conti *et al.*^50^ (Methods: *Formulating Feature Sets*). We also constructed an independent test set from a subset of the UKBB to fairly assess model generalization (Methods: *Datasets*).

To contextualize VADEr’s performance we optimized a number of baseline models including classical C+T, LDpred2^30^, and five different ML approaches (Methods: *Baselines*). We also reconstructed the 269 variant PCa PRS reported by Conti *et al.*^50^, the study from which we acquired our summary statistics. As detailed in Methods: *Baselines*, VADEr and the ML baselines must learn to map genomic features to labels exclusively from the training set (n = 73,315), while the remaining PRS methods (i.e., C+T, LDpred2, and the 269 Conti *et al.* PRS) can leverage a much larger effective sample size (n = 234,253) by using effect sizes from the original Conti *et al.* summary statistics. To ensure a fair comparison across models, we also generated training set specific C+T and LDpred2 baseline PRSs, using effect sizes derived exclusively from the training set. Including both provides a contextualized and fair assessment of VADEr’s performance relative to equivalent and optimized (indicated by ^➕^) summary-statistic-based baselines.

After training, all models were applied to the held out test set of UKBB samples. VADEr outperformed all statistical genomics baselines (including the Conti *et al.* PRS). Specifically, compared to LDpred2^➕^, the top-performing statistical genomics baseline across metrics, VADEr achieved absolute increases of 0.007 in AUC, 0.243% in accuracy, 0.008 in average precision (AP), 0.016 in F1 score, and 0.006 in Matthew’s correlation coefficient (MCC). These differences double when limiting statistical genomics models to training set specific effect sizes (**Table 2**). Against the ML baselines, VADEr emerged as the top-performing model across 3 of the 5 metrics, including accuracy, AP, and MCC, with particularly strong gains in accuracy (1.03%) and MCC (0.008). AP gains were more modest (0.0003), with XGB and logistic regression showing comparable performance. VADEr also ranked second in AUC, with performance closely matching that of logistic regression, the leading model by this metric. We note the difference in AUC between VADEr and logistic regression was statistically insignificant with a DeLong test^66^ (p = 0.17). Overall, VADEr demonstrated the best performance in accuracy, AP, and MCC, and ranked a close second in AUC. A detailed breakdown of model performances across metrics can be found in **Table 2**.

### Linking VADEr Attributable Risk to Loci With DARTH Scores

In order to determine whether VADEr indeed captured risk-relevant information from complex, non-linear relationships embedded within the underlying genomic data, we developed **D**irected **A**ttention **R**elevance from **T**ransformer **H**euristics (DARTH) scores. Based on an extension of Beyond Attention introduced by *Chefer et al.*,^61,63^ DARTH scores represent VADEr’s patch-specific attentional attribution fingerprint, and serve as a metric of each region’s influence towards driving disease risk for an individual. This approach provides a personalized interpretability metric grounded in the model’s internal attention patterns, that addresses limitations of classical attention-based interpretability methods and clarifies how VADEr attributes and integrates genomic signals to inform disease risk (Methods: *DARTH Patch Attribution*).

To evaluate population-level patch contributions to PCa risk predictions, we averaged DARTH patch contributions across a dataset (e.g., the test set) to obtain a metric of global model attributed risk. Comparing these averaged attributions with variant significances reported in the GWAS summary statistics generated by Conti *et al.*, we found that VADEr’s highest attributable risk regions generally aligned with those containing the most significant GWAS variants, though specific differences were observed in both the ELLIPSE validation set (**Supplementary Figure 1**) and the UKBB test set (**Figure 2A**). For example, VADEr’s top three global attributable regions of risk fell across consecutive patches in the chromosome 8q24 region, a well-documented locus for PCa risk^67,68^ (**Figure 2B**). Additionally, high risk patches were detected on chromosomes 10 and 17. The chromosome 10 top risk patch (10q11.2) encompasses both the *MSMB* gene and the documented PCa risk variant rs10993994,^69,70^ which modulates *MSMB* expression, one of the most abundant proteins secreted by the prostate.^71^ Chromosome 17 exhibited multiple high-attribution patches that differed in distribution from GWAS signals, suggesting alternative risk localization patterns.

**Figure 2:**
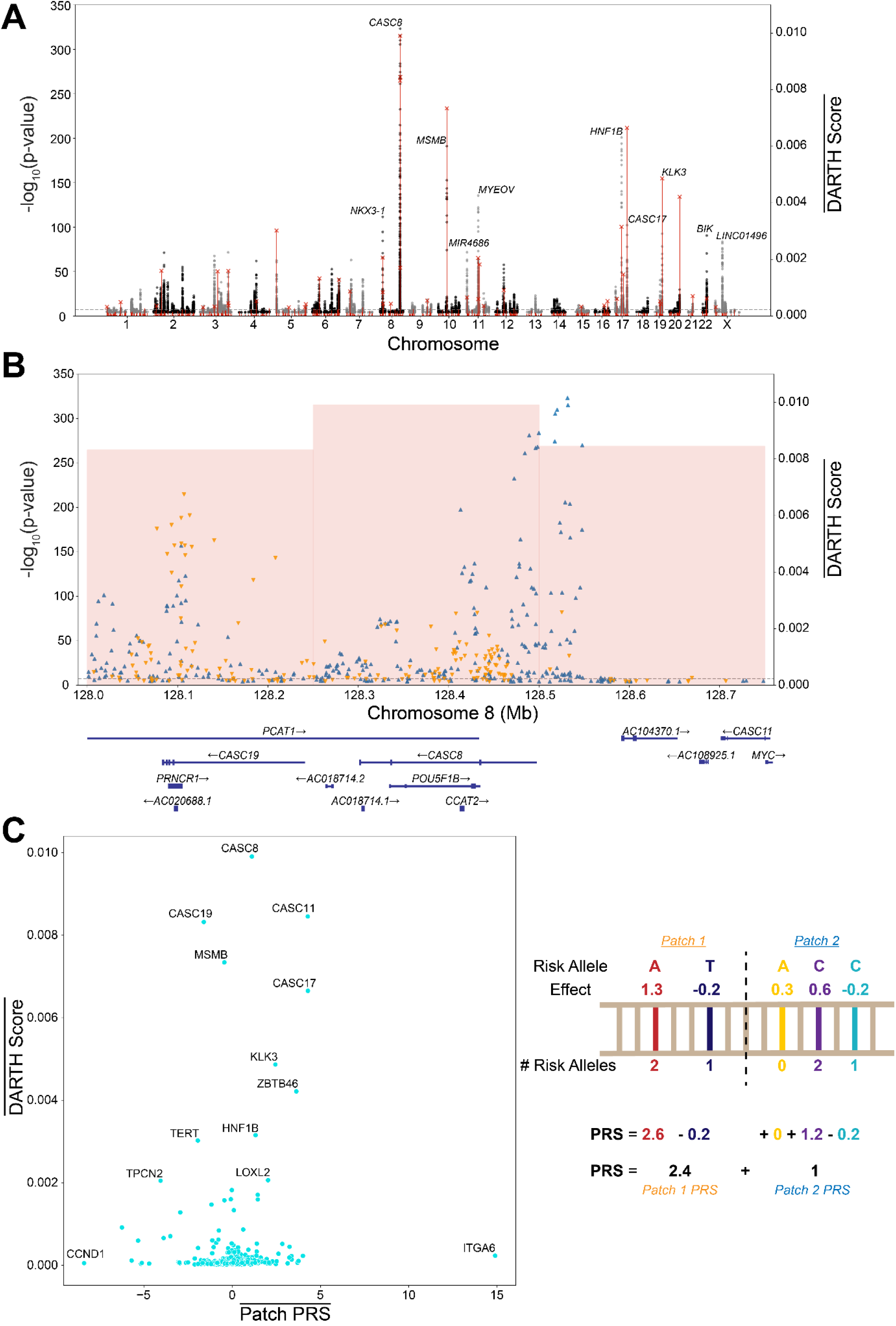
Comparing patch average attribution alignment between classical PRS and VADEr with DARTH scores. A) DARTH Manhattan plot: Per patch average DARTH scores (red lines) are superimposed on summary statistic p-values from Conti *et al.* (black and gray circles). The dotted horizontal line corresponds to the genome-wide significance p-value of 5 × 10^−8^. B) Close-up of chromosome 8 risk region. Three contiguous patches on chromosome 8 spanning from 128-128.75 Mb overlay with the largest p-value peak containing regions encoding the oncogene *MYC* and PCa associated lncRNAs PCAT1, PRNCR1, CASC8, CASC11, and CASC19. Triangle orientation for variants indicates direction of risk effect size (i.e., orange variants indicate a protective PCa effect and blue variants a predisposing PCa effect). C) Assessing average VADEr attributions through DARTH scores against the average patch contributions to a classical additive PRS. Labeled regions correspond to genes found in a given patch.

To further contextualize VADEr’s attributions, we compared DARTH scores with patch-level risk attributed by a classical (weighted genotype summation) PRS formulation employing the same subset of features as VADEr and effect sizes from Conti *et al.*^50^ As with DARTH scores, patch-level classical PRS contributions were averaged across the test set to yield a comparable risk profile (**Figure 2C**). While we observed notable areas of concordance, such as patches on chromosome 8, 17, 19, and 20 receiving high attributed risk across approaches, divergences also emerged. For instance, the *ITGA6* patch (starting at chromosome 2:173.25 Mb and containing 73 SNPs) was the highest risk region globally for the classical PRS, but ranked 60th in VADEr’s attributions, indicating a difference in relative prioritization between models. A natural assumption might be that the contributions of risk at this region effectively serve as a bias for the classical PRS, simply additively shifting the score distribution. However, we find this not to be the case, as this patch exhibits the ninth highest classical PRS patch variability (*σ*^2^ = 3.66) across the entire test set, just below the top three chromosome 8 patches. It might next be assumed that the magnitude of classical PRS risk for this patch is attributable to the patch density (i.e., the number of SNPs in the patch). However, we find that this region falls outside of the top 20 patches in terms of patch density. Given discrepancies between top attributed patches between approaches, we questioned whether DARTH scores could be used to constrain the feature space of a classical PRS and thus improve performance. Despite differences in methodology, with VADEr’s architecture geared towards capturing complex non-linear interactions, we found that reducing the number of patches used by a classical additive PRS to the top 25% and 50% of patches with the highest average DARTH score yielded improvements in stratification potential by AUC, with gains of 0.27% and 0.84% respectively.

Given the variable SNP density across patches, we next investigated the relationship between patch density and averaged DARTH scores. While these variables exhibited a moderate correlation (Pearson r = 0.51), we observed pronounced exceptions to this phenomenon, most notably, with two single SNP patches falling among the top 15 average DARTH scores. To our knowledge, neither of these SNPs, rs13013607 (Conti *et al.* GWAS p = 7.59 × 10^−6^), located on chromosome 2 near *CALM2*, *STPG4*, and *TTC7A*, nor rs1624540 (Conti *et al.* GWAS p = 3.13 × 10^−5^), an intron variant in *INTS4* on chromosome 11, is currently a documented PCa risk variant. However, their high average attribution by VADEr suggests their potential for further investigation, particularly in European populations. We note that rs13013607 is over 125 Mb away from the *ITGA6* patch, the top chromosome 2 classical PRS patch highlighted above (**Figure 2C**), indicating that its high average DARTH score is independent of variant correlation between these patches.

While averaged attributions offer insights into global risk patterns and method concordance, they obscure individualized patch contributions to disease risk. To assess VADEr’s ability to capture genetic heterogeneity contributions to disease risk, we employed UMAP reduction^72^ to project the 1204-dimensional DARTH manifold to two dimensions, transforming each test set individual into a distinct two-dimensional point. This projection revealed a clear gradient of VADEr prediction values across the dimensions, with a dense central cluster displaying less defined substructure and three distinct outlying clusters, suggestive of groups of individuals with divergent genetic contributions driving disease risk (**Figure 3A**). Indeed, an investigation into patch attributions driving these three outlier clusters revealed specific patches, overlapping *HOXB13*, *TMPRSS2*, and *MSMB* respectively, driving predicted disease risk. A detailed analysis of these findings is presented below, with each cluster examined in its respective subsection.

**Figure 3:**
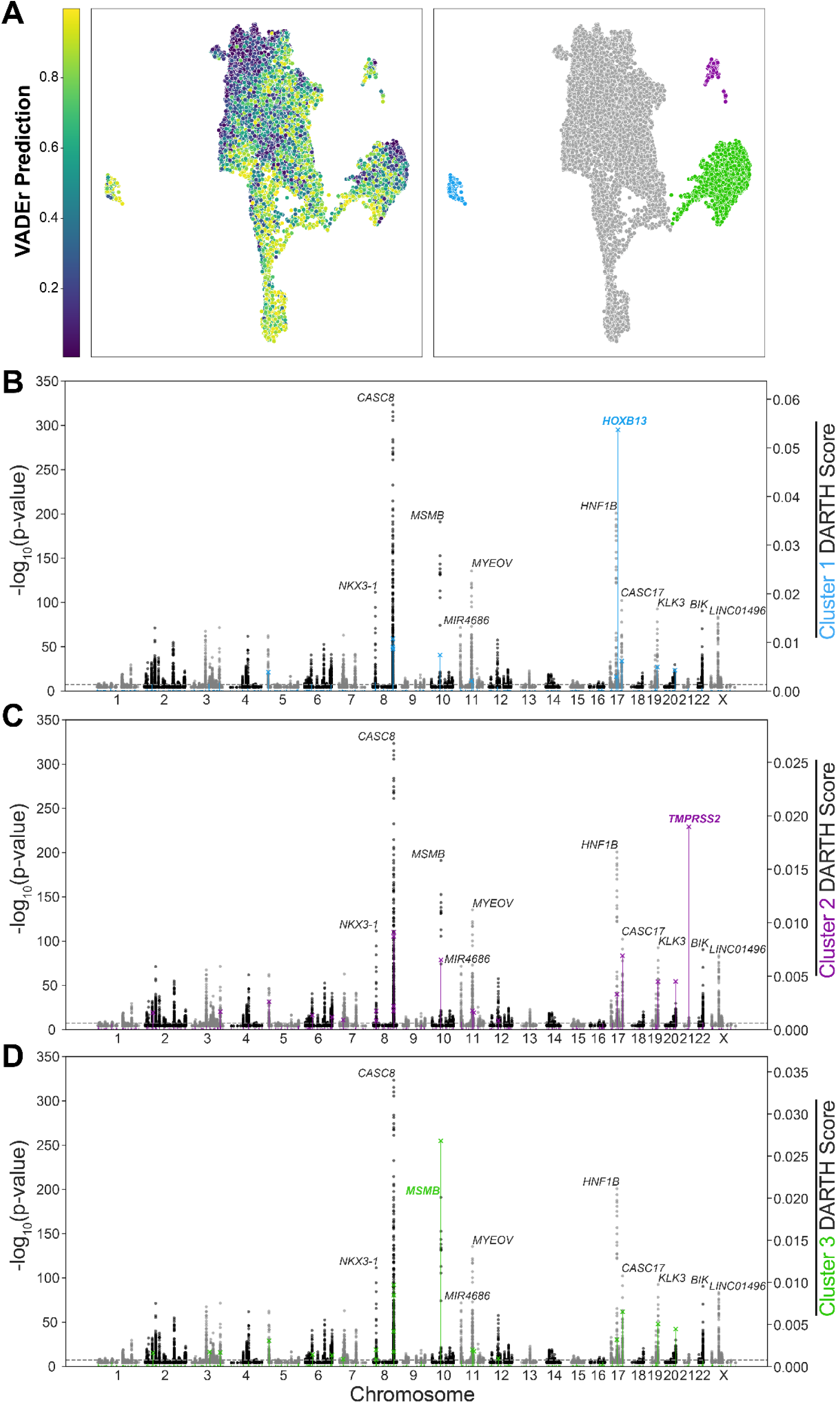
Identifying genetic substructure driving predicted risk from DARTH scores. A) UMAP projection of test set individuals using patch DARTH scores. Shading by VADEr prediction values show a clear gradient across the dimensions (left) with three distinct outlying clusters (right). B) Across individuals in cluster 1 (light blue), the DARTH score distribution deviates from the global average test set distribution (Figure 2A), with the patch spanning 46.75-47 Mb on chromosome 17 containing 11 variants dominating risk attribution. C) Cluster 2 (purple) exhibits a similar phenomenon with the patch spanning 42.75-43 Mb on chromosome 21 containing 30 variants identified as the dominant region of relevance. D) The dominant driver of predicted risk in cluster 3 (green) individuals aligns with the chromosome 10 tower spanning 51.5-51.75 Mb containing 31 variants and encompassing the gene *MSMB*.

#### Cluster 1 (n = 317): *HOXB13* Region

Individuals in cluster 1 exhibited a pronounced elevation in DARTH score for the patch starting at chromosome 17:46.75 Mb, the region which encodes for *HOXB13* (**Figure 3B**). This patch, which ranked 17th among all patches by global average DARTH score and 3rd among all patches on chromosome 17, exhibited the highest DARTH score bimodality coefficient (BC) of any patch in the test set (BC = 0.90), and was distinctly uniquely elevated amongst individuals in this cluster (**Supplementary Figure 2A**). Notably, the variant rs138213197, corresponding to a HOXB13 G84E mutation, is known to significantly increase PCa risk in individuals of European descent,^35,36^ particularly in Nordic populations.^73^ Though this variant was absent from the feature set, VADEr may have inferred its presence indirectly. Across the test set, 126 individuals were identified as carriers of this variant (all heterozygous), with 107 of these individuals found within cluster 1. Regardless, VADEr captured heightened PCa risk from the variants in this patch among individuals in this cluster (OR = 1.75; p = 1.52 × 10^−6^) relative to those in the dense central cluster (n = 12,026). Cluster 1 individuals also showed a significant increase in identity by state (IBS) at this patch (mean patch IBS = 0.83) relative to a matched size random sampling of non-cluster 1 individuals from the test set (mean IBS = 0.76) consistent with ancestry-associated risk.

#### Cluster 2 (n = 303): *TMPRSS2* Region

Among individuals in cluster 2, the patch starting at chromosome 21:42.75 Mb, the region encoding *TMPRSS2*, displayed a markedly higher DARTH score relative to other patches (**Figure 3C**). Similar to the dominant patch in cluster 1, this patch, which was ranked 23rd among all patches by global average DARTH score, exhibited the second highest bimodality of any patch in the test set (BC = 0.88), and was predominantly elevated amongst individuals in this cluster (**Supplementary Figure 2B**).

A defining molecular subtype in PCa is the presence of a TMPRSS2-ETS gene fusion, predominantly involving ERG (though ETV1 and ETV4 are found to a lesser extent).^37,38^ This fusion, which is found in ∼50% of PCa cases, is a significant driver in the etiology of PCa, as it results in the constitutive expression of oncogenic ETS factors.^38,74^ That VADEr implicated the *TMPRSS2* locus as a germline risk region for PCa raises the possibility that haplotypes at this locus may facilitate fusion formation, though it is also possible that haplotypes here may contribute to PCa risk through fusion-independent mechanisms.

Unlike cluster 1 where the *HOXB13* patch had a fivefold increase in mean cluster DARTH score (**Figure 3B**), cluster 2 displayed a more distinct pattern of DARTH scores among patches. The dominant *TMPRSS2* patch in cluster 2 showed only a two-fold increase in mean cluster DARTH score relative to the next highest attributed patch (**Figure 3C**). Other significant patches in cluster 2 included those identified on chromosomes 8, 10, and 17. In relation to the TMPRSS2-ERG fusion-positive PCa subtype, SNPs in each of these patches, in particular rs16901979 (8q24), rs1859962 (17q24), and the aforementioned rs10993994 (10q11) have all been shown to be associated with differences in the presence of this subtype.^75^ rs1859962 is located in the coding region of CASC17 and is part of the feature set, within the top identified global average chromosome 17 patch (**Figure 2A**). Although rs16901979 was not in the feature set, it is possible that the risk attributed to this SNP was captured by the patch representations of the previously mentioned contiguous chromosome 8 patches with the highest global average VADEr attribution (**Figure 2B**). Taken together, this cluster’s DARTH score signature suggests a collective germline contribution capable of influencing TMPRSS2-ETS gene fusions during PCa progression.

Cluster 2 individuals exhibited increased IBS at the *TMPRSS2* patch (mean patch IBS = 0.84) compared to a a matched size random sampling of non-cluster 2 individuals (mean patch IBS = 0.79), though genome-wide IBS was equivalent across both groups (mean IBS = 0.74). Individuals in cluster 2 also demonstrated significantly elevated risk compared to the dense central cluster (OR = 1.34; p = 0.012).

#### Cluster 3 (n = 2,589): *MSMB* Region

Unlike clusters 1 and 2, for which the dominating DARTH score patches were regions underemphasized or otherwise overlooked by a global assessment, the key identified driver of risk for individuals in cluster 3 was the patch beginning at chromosome 10:51.5 Mb (**Figure 3D**). This patch, which was the top chromosome 10 risk region and 4th highest risk region overall in our global analysis (**Figure 2A**), includes, as previously mentioned, both the *MSMB* gene and the well-documented PCa risk variant rs10993994. This is notable given that the protein for which *MSMB* encodes exhibits broad tumor suppressive properties,^76^ and rs10993994 is a documented modulator of this gene’s expression.^77^ Though rs10993994 is in this patch, it is unlikely that the emphasis of effect attributed to this region is driven solely by contributions of this SNP. There are 31 variants encoded in this patch, 8 of which fall within the coding region of *MSMB*, and 3 (including rs10993994) positioned less than 1500 bp upstream of this gene. Several of these SNPs have been investigated for their association with *MSMB* expression,^78^ suggesting that multiple variants in this region may contribute to PCa risk through effects on *MSMB* regulation. Moreover, as with clusters 1 and 2, a significantly higher (p ≪ 0.001) mean IBS for this patch (mean patch IBS = 0.87) was observed relative to a matched size random sampling of non-cluster individuals (mean patch IBS = 0.78). This, in conjunction with consistent genome-wide similarity between groups (mean IBS = 0.74), indicates a region-specific contribution to attributed predicted risk by VADEr. Individuals in this cluster, like those in clusters 1 and 2, also exhibited significantly elevated risk relative to the dense central cluster (OR = 1.36; p = 1.32 × 10^−12^).

As expected, given the *MSMB* patch’s high global average DARTH score and relatively low bimodality (BC = 0.47) compared to the top patches identified in clusters 1 and 2, its DARTH score cluster signature is less distinct (**Supplementary Figure 2C**). Nonetheless, within cluster 3, the *MSMB* patch showed a three-fold increase in mean cluster DARTH score relative to the next highest attributed patch (**Figure 3D**). This pronounced difference emphasizes the unique contribution of the *MSMB* region to risk in cluster 3 individuals, particularly in contrast to the dense central cluster, where attributed risk is primarily concentrated by the contiguous chromosome 8 patches spanning 128-128.75 Mb (**Supplementary Figure 2D-F**).

### Empirical Validation of DARTH Scores and Regional Risk Attribution

While DARTH scores represent a metric of personalized patch-level attribution capable of identifying heterogeneous genetic contributions to disease, their alignment with predicted risk has yet to be empirically demonstrated. Additionally, it remains unclear whether outlier patch-driven cluster signatures arise from predominantly regional contributions, as captured by patch-level embeddings, or from broader genomic context dependencies. The inflation in patch-level IBS scores for cluster-specific outlier patches suggests, though does not confirm a regional effect.

To address these questions, we conducted a patch perturbation analysis in the UKBB (Methods: *Perturbing Patches*). Comparison of the prediction difference distributions between inter- and intra-cluster patch shuffling supports a locus-driven risk hypothesis, suggesting VADEr’s emphasis on outlier patches is attributable to its patch-level embeddings. Among cluster 1 and its matched size random complement, we observed minimal variation in PCa prediction probability when performing intra-cluster shuffling for the dominant cluster 1 *HOXB13* patch. However, inter-cluster shuffling of this patch led to significant shifts in predicted risk for both groups, with cluster 1 individuals having a reduced median predicted risk of PCa by 13.79% (p = 4.73 × 10^−102^) and random complement individuals having an increased median predicted risk of PCa by 13.52% (p = 2.29 × 10^−101^; **Supplementary Figure 3A**). No other patches exhibited a significant distribution deviation in VADEr prediction probability difference between the two shuffling methodologies in either group.

Similarly, for cluster 2, inter-cluster shuffling relative to intra-cluster shuffling of the dominant *TMPRSS2* risk patch significantly reduced predicted median risk of PCa in cluster individuals by 8.97% (p = 4.77 × 10^−89^) and significantly increased it in random complement individuals by 7.51% (p = 2.52 × 10^−78^; **Supplementary Figure 3B**). Likewise, in cluster 3, inter-cluster shuffling of the dominant *MSMB* risk patch led to a significant 8.35% (p ≪ 0.001) decrease in predicted median risk in cluster individuals and a significant 8.10% (p ≪ 0.001) increase in corresponding random complement individuals (**Supplementary Figure 3C**). Modest though significant distribution deviations were also observed for the CASC19 containing patch in both cluster 2 and 3 (**Supplementary Figure 3B,C**). Across all analyses, we observed that variability in VADEr prediction probability difference, regardless of shuffling methodology, tracked with global average DARTH attribution scores (**Supplementary Figure 3**), further supporting the ability of DARTH scores to capture personalized contributions to disease risk.

### DARTH Score Signatures Reveal Germline Predisposition for Molecular Subtypes

VADEr’s heterogeneous patterns of germline risk attribution, as revealed through DARTH scores, highlight a natural avenue for further exploration: whether distinct model-derived risk signatures align with known molecular subtypes of PCa. To investigate this, we obtained germline genotypes and molecular subtype annotations for European ancestry PCa individuals in TCGA (Methods: *Datasets*). Genotype imputation allowed recovery of 11,149 of the 12,017 variants in the VADEr feature set, with the remaining missing variant genotypes assigned according to the median SNP values from the UKBB.

Prior to conducting DARTH score-subtype associations, we assessed VADEr’s prediction and attribution robustness in TCGA. Encouragingly, VADEr predictions were significantly inversely correlated with age of diagnosis in the case-only dataset (Pearson r = −0.143; p = 0.004). We found no evidence of dataset-specific batch effects in DARTH scores between TCGA and UKBB, and observed the same cluster risk signatures in TCGA individuals as previously identified in the UKBB (**Supplementary Figure 4**). Among the 8 TCGA individuals exhibiting a *HOXB13* cluster signature, all G84E mutation variant (rs138213197) carriers in TCGA (n = 6), were found in this cluster. Independent of cluster signatures, we also observed that, similar to the UKBB, the single SNP patches containing rs13013607 and rs1624540 were among the 15 highest attribution patches by average DARTH score in TCGA, providing further evidence in support of a potential role of these variants in PCa risk.

To identify DARTH score associations with molecular subtypes, we proceeded with the subset of TCGA PCa individuals for which subtype annotations were available (n = 275/412). These subtypes, based on particular oncogenic drivers,^39^ spanned seven classes including ETS gene fusions in: 1) *ERG* (n = 137), 2) *ETV1* (n = 24), 3) *ETV4* (n = 11), or 4) *FLI1* (n = 3); or mutations in 5) *SPOP* (n = 25), 6) *FOXA1* (n = 8), or 7) *IDH1* (n = 2). The remaining individuals (n = 65) were not associated with a defined subtype and were grouped together in an eighth *Other* class. We note that the four defined ETS fusion subtypes are not specific to fusions with *TMPRSS2*, and can include fusion events with other androgen-related genes including *SLC45A3* and *NDRG1*.^39^ Given this, we constructed a ninth, non mutually-exclusive *TMPRSS2-ETS* subtype specific to TMPRSS2-ETS gene fusions (n = 131).

For the *SPOP* subtype, we observed a single significant DARTH score association after multiple-hypothesis correction with the patch starting at chromosome 7:97.75 Mb (point-biserial correlation r_pb_ = 0.30, p = 4.70 × 10^−4^; AUC = 0.563; AP = 0.274). Permutation testing (Methods: *Statistical Analyses*) provided further support for this region’s relevance to the *SPOP* subtype, with observed correlation and AP values significantly exceeding those obtained under an empirical null distribution generated by random label permutations (p_r_ = 0.001; p_AP_ = 0.001). This patch, which encodes for 5 protein coding genes, includes *LMTK2*, a gene well-documented for its association with PCa risk^79^ and known to negatively regulate androgen receptor (AR) transcriptional activity,^79,80^ a signaling pathway central to growth and survival programs in PCa.^39^ Wild-type *SPOP* plays a similar tumor suppressor role in PCa, inhibiting AR transcriptional activity.^81^ When *SPOP* mutations occur however, this function is lost, and significantly elevated AR activity has been observed, including in the TCGA *SPOP* subtype investigated here.^39,81^ Taken together, this suggests a plausible path to germline *SPOP* subtype predisposition as identified by VADEr. Specifically, germline variants or haplotypes at this region may detrimentally affect *LMTK2*, through reduced expression or function, weakening its ability to suppress AR activity. This in turn would create a cellular context in which tumorigenic processes that further elevate AR signaling, such as *SPOP* mutations, are selectively favored, and ultimately, bias tumor evolution towards this subtype.

For the *FLI1* and *IDH1* subtypes, we identified 2 and 6 patches, respectively, with significant DARTH score-subtype associations and corroborated with permutation testing. Interestingly, the cluster 2 signature *TMPRSS2* patch was among the 6 patches associated with the *IDH1* subtype, which is notably ETS-fusion negative, and instead defined by an oncogenic hotspot mutation at R132 and genome-wide DNA hypermethylation.^39,82^ However, the overall sparsity of both the *IDH1* and *FLI1* subtypes in our cohort warrants caution in interpreting these findings and underscores the need for validation in larger, independent datasets with paired tumor and normal genomic data. A detailed summary of all significant DARTH score-subtype associations can be found in **Supplementary Table 1**.

No significant associations were observed between DARTH scores and the *ERG*, *ETV1*, *ETV4*, *TMPRSS2-ETS*, *FOXA1*, or *Other* subtypes. Notably, none of the aforementioned SNPs (rs16901979, rs1859962, and rs10993994), previously reported^75^ to have been associated with the presence of TMPRSS2-ERG fusion PCa, associated with the subtype here. This may reflect limited power to detect modest effects given the size and composition of the dataset, and thus suggests further investigation may be warranted.

## Discussion

In this work, we introduce VADEr, a novel ViT-inspired architecture that integrates techniques from both NLP and CV to align with the unique characteristics of genomic data, capture interactive effects, and enhance predictive power in genotype-to-phenotype prediction. As proof-of-concept, we assessed its performance in PCa prediction, and observed that across a range of metrics, including accuracy, AP, and MCC, VADEr outperformed other polygenic risk prediction methods, even those leveraging a much larger effective sample size through use of external summary statistics (**Table 2**).

An investigation of baselines by PRS methodology (i.e., summary-statistic-based vs. ML-based) reveals an interesting, if perhaps not unexpected, finding. Specifically, most ML-based PRS approaches showed potential to yield predictive improvements over their summary-statistic-based counterparts, particularly for rank-based metrics such as AUC and AP. However, unlike VADEr, this improvement was not universal across metrics, as LDpred2^➕^ outperformed ML baselines globally in accuracy and MCC. Despite this, it is important to highlight the potential of appropriately regularized and hyperparameter optimized ML methods, even if simplistic, to improve genotype-to-phenotype prediction performance, especially if trained on datasets of size comparable to those from which large-scale summary statistics are computed.

Although VADEr outperformed baseline methods across several metrics, the gains might be perceived as modest. Given this, it is important to contextualize these improvements within the inherent limitations of complex disease heritability. In this study we investigated prostate cancer (PCa), a condition for which heritability is estimated to be approximately 57%.^83,84^ Though this estimate indicates a substantial genetic contribution, it also highlights an upper bound on the predictive power of what is achievable through genetic data alone. Thus, given the limitations imposed by disease heritability, even incremental performance gains represent meaningful advancements, and further emphasize that we are working at the margins of what is genetically predictable.

AUC is a commonly used, rank-based metric for assessing the performance of PRS models.^2,10^ Interestingly, we found logistic regression to outperform VADEr by this metric, though not significantly (p = 0.17). However, it is important to consider the paradoxical relationship that can exist between desirable model attributes, such as identifying distinct genetic heterogeneity, and their potential to deflate performance on a metric such as AUC. For example, VADEr identified the high risk DARTH score dominant *HOXB13* and *TMPRSS2* clusters, both of which exhibited an enrichment in predicted cases, which by definition corresponds to elevated prediction probabilities. Though this stratification enhances the identification of high-risk individuals, it also introduces a cluster-shifting effect, which effectively concentrates high prediction scores in a way that can reduce overall rank differentiation. This highlights the limitations of relying solely on rank-based metrics when evaluating models capturing complex genetic substructures, and emphasizes the need for a multifaceted assessment of model performance.

Independent of rank-based metric intricacies, it is noteworthy that VADEr achieved strong performance despite the limited training set size relative to the model’s complexity. With a 12-layer, 12-headed Transformer architecture and a 768-dimensional embedding space, the number of trainable model parameters (n = 96M) far exceeds the number of training samples (n = 73,315) and features (n = 12,017). Drawing parallels to the original ViT study, which found that Transformers can underperform convolutional neural networks (CNNs) on mid-sized datasets without strong regularization or large-scale pre-training,^12^ our findings suggest that VADEr is well-suited to genotype-to-phenotype modeling even without extensive data, augmentations, or pre-training. Moreover, it is reasonable to anticipate that as more genomic data become available, allowing for increased training set sizes, the magnitude of VADEr’s performance gains could increase, though underlying disease heritability naturally imposes an upper bound.

While VADEr and other ML-based methods afford avenues for improving PRS performance, these approaches are of limited practical and clinical utility without a means to identify, interpret, and validate factors driving prediction. To that end, we complement the VADEr architecture with a local (i.e., individual-specific), region-level interpretability metric, DARTH scores. These scores, which leverage attention-based attribution techniques introduced by Chefer *et al*.,^61,63^ when collapsed to the global average resolution exhibit a pattern of attributed risk that closely, though not identically aligns with GWAS-identified high-risk regions (**Figure 2A**; **Supplementary Figure 1**). Specifically, well-documented PCa risk regions at 8q24, 10q11, and 17q24 were all identified as significant drivers of predicted risk by VADEr across the test set. Additionally, VADEr identified, in both the UKBB and TCGA, two globally prominent single-SNP patches on chromosomes 2 (rs13013607) and 11 (rs1624540), which to our knowledge have not yet been documented as PCa risk variants. This suggests these SNPs, or the regions in which they fall, may represent novel drivers of PCa risk in individuals of European descent.

By leveraging the inherent personalized properties of DARTH scores, we gained unique mechanistic insights into VADEr’s attributed architecture of genomic risk. Across the test set, we observed three distinct clusters driven by risk patterns that notably deviated from both the global average and GWAS-level perspectives (**Figure 3**). Each cluster exhibited an outlier dominated signature, with a specific patch dominating the attribution profile. While each cluster signature highlights a biologically relevant dominant risk region, cluster 2 (n = 303) warrants particular attention for its *TMPRSS2* signature. In contrast to the identified risk loci of *HOXB13* and *MSMB* in clusters 1 (n = 317) and 3 (n = 2,589) respectively, for which germline risk for PCa is well-documented, *TMPRSS2* is mainly known for its eponymous role in TMPRSS2-ERG fusion-positive PCa, with its region’s germline contributions to PCa risk far less characterized. Given this, our identification of a *TMPRSS2*-dominant risk signature is particularly noteworthy, especially when considered among other emphasized regions in the signature. Specifically, cluster 2 also highlighted regions containing SNPs previously associated with the TMPRSS2-ERG subtype (e.g., rs16901979 at 8q24, rs1859962 at 17q24, rs10993994 at 10q11).^75^ However, to our knowledge, no SNPs within the *TMPRSS2* locus itself have been documented in this context. Taken together, these findings suggest that the germline architecture of the *TMPRSS2* region may not only contribute to overall PCa risk, but also shape the pathogenesis of this fusion-driven subtype. While we were unable to validate this in TCGA, it remains an avenue that warrants further exploration.

Beyond providing mechanistic insight into individualized VADEr-identified genomic risk, DARTH scores afford advantages that extend beyond general disease risk attribution. Though VADEr training, inference, and by extension DARTH score formulation, are independent of molecular subtype annotations, DARTH score distributional patterns across individuals were capable of revealing germline associations with PCa molecular subtypes. Most notably, we identified a predisposition for the *SPOP* molecular subtype with elevated DARTH scores at the *LMTK2* region (chromosome 7:97.75 - 98 Mb). Given the shared tumor-suppressive role of wild-type *SPOP* and *LMTK2* in regulating AR transcriptional activity,^79–81^ this association suggests a plausible germline-driven mechanism for subtype-specific risk. Overall, the ability of DARTH scores to identify regions of germline risk with molecular subtypes, without explicit supervision, has significant implications for personalized risk stratification, biomarker discovery, and therapeutic targeting. Nevertheless, further investigation and validation of DARTH score-subtype associations are needed to fully assess their potential and clinical utility.

In conclusion, our study demonstrates the efficacy of the VADEr architecture as a powerful approach for genotype-to-phenotype modeling, particularly in the context of complex diseases like PCa. Coupled with our introduction of DARTH scores, a robust, personalized, and region-specific interpretability metric, we afford a means to elucidate VADEr identified genetic substructure. However, it is important to reiterate the proof-of-concept nature of this work and highlight several practical considerations, especially those that pertain to portability, that require addressing. Our study focused primarily on PCa prediction in European-ancestry individuals and leveraged external summary statistics for feature selection, which, despite our best efforts, may influence true model generalizability. To address this future investigations should expand upon our work, ensuring VADEr’s broad applicability in further independent cohorts, diverse populations, and other genotype-to-phenotype contexts. Moreover, assessing and addressing model robustness in the face of missing genomic features is critical for practical adoption, and underscores the need for future architectural adaptations and training procedures to enable reliable inference with incomplete data. Finally, extending VADEr’s framework to incorporate rare, high-impact variants (e.g., BRCA2), either directly as model inputs or indirectly in conjunction with VADEr-derived risk predictions, represents a promising avenue for future improvement. Overall though, by advancing our understanding of genetic architectures underlying complex diseases, DARTH-VADEr offers a promising path towards improving both genomic risk scores and personalized medicine.

## Author Contributions

Original concept, J.V.T.; project supervision, H.C.; VADEr development and training, J.V.T.; baseline modeling, J.V.T., A.K., G.P.; DARTH-VADEr interpretability, J.V.T.; data acquisition and processing, J.V.T., A.K., M.S.P.; prostate cancer interpretation advising, B.R., T.S.; polygenic risk score advising, M.G.; preparation of manuscript, J.V.T., H.C.

## Data and Code Availability

All data utilized in this study were obtained from public sources. Specifically, ELLIPSE genotypes and phenotypes were downloaded from dbGaP (study accession: phs001120.v1.p1). UKBB data was retrieved under project ID 37671. TCGA data was obtained from the TCGA Genomic Data Commons Data Portal. The complete codebase for this study, including the full VADEr model architecture and tailored implemented training procedures, can be found on our GitHub at https://github.com/jvtalwar/DARTH_VADEr.

## Acknowledgements

The results presented here are in large part made possible by data generated by the ELLIPSE Consortium (dbGaP study: phs001120), TCGA Research Network (https://www.cancer.gov/tcga), and the UKBB (Project #37671). This work was supported by Emerging Leader Award from The Mark Foundation for Cancer Research, grant #18-022-ELA, NIH Grant R01CA269919 to H.C., and infrastructure grant 2P41GM103504-11.

For the dbGaP study phs001120 we acknowledge: Funding for the meta-analysis was provided by NIH grant U19CA148537. For de novo genotyping we would like to acknowledge the NCRN nurses and consultants for their work in the UKGPCS study. We thank all the patients who took part in this study. This work was supported by Cancer Research UK (grant numbers C5047/A7357, C1287/A10118, C1287/A5260, C5047/A3354, C5047/A10692, C16913/A6135 and C16913/A6835). We would also like to thank the following for funding support: Prostate Research Campaign UK (now Prostate Cancer UK), The Institute of Cancer Research and The Everyman Campaign, The National Cancer Research Network UK, The National Cancer Research Institute (NCRI) UK. We are grateful for support of NIHR funding to the NIHR Biomedical Research Centre at The Institute of Cancer Research and The Royal Marsden NHS Foundation Trust. The MEC was supported by NIH grants CA63464, CA54281 and CA098758.

## References

1. Torkamani, A., Wineinger, N. E. & Topol, E. J. The personal and clinical utility of polygenic risk scores. Nat. Rev. Genet. 19, 581–590 (2018).

2. Lewis, C. M. & Vassos, E. Polygenic risk scores: from research tools to clinical instruments. Genome Med. 12, 44 (2020).

3. Devlin, J., Chang, M.-W., Lee, K. & Toutanova, K. BERT: Pre-training of deep bidirectional Transformers for language understanding. Proceedings of NAACL-HLT (2019) doi:10.18653/v1/N19-1423.

4. Gasperini, M., Tome, J. M. & Shendure, J. Towards a comprehensive catalogue of validated and target-linked human enhancers. Nat. Rev. Genet. 21, 292–310 (2020).

5. Avsec, Ž., et al. Effective gene expression prediction from sequence by integrating long-range interactions. Nat. Methods 18, 1196–1203 (2021).

6. Pombo, A. & Dillon, N. Three-dimensional genome architecture: players and mechanisms. Nat. Rev. Mol. Cell Biol. 16, 245–257 (2015).

7. Wu, H. et al. CvT: Introducing convolutions to vision transformers. in 2021 IEEE/CVF International Conference on Computer Vision (ICCV) (IEEE, 2021). doi:10.1109/iccv48922.2021.00009.

8. Slatkin, M. Linkage disequilibrium--understanding the evolutionary past and mapping the medical future. Nat. Rev. Genet. 9, 477–485 (2008).

9. Vilhjálmsson, B. J. et al. Modeling Linkage Disequilibrium Increases Accuracy of Polygenic Risk Scores. Am. J. Hum. Genet. 97, 576–592 (2015).

10. Choi, S. W., Mak, T. S.-H. & O’Reilly, P. F. Tutorial: a guide to performing polygenic risk score analyses. Nat. Protoc. 15, 2759–2772 (2020).

11. Vaswani, A. et al. Attention is All you Need. in Advances in Neural Information Processing Systems (eds. Guyon, I. et al.) vol. 30 (Curran Associates, Inc., 2017).

12. Dosovitskiy, A., et al. An image is worth 16×16 words: Transformers for image recognition at scale. ICLR (2021).

13. Brown, T. B. et al. Language Models are Few-Shot Learners. arXiv [cs.CL] (2020).

14. Caron, M. et al. Emerging properties in self-supervised vision transformers. in 2021 IEEE/CVF International Conference on Computer Vision (ICCV) 9630–9640 (IEEE, 2021).

15. OpenAI et al. GPT-4 Technical Report. arXiv [cs.CL] (2023).

16. Ji, Y., Zhou, Z., Liu, H. & Davuluri, R. V. DNABERT: pre-trained Bidirectional Encoder Representations from Transformers model for DNA-language in genome. Bioinformatics 37, 2112–2120 (2021).

17. Zhou, Z. et al. DNABERT-2: Efficient foundation model and benchmark for multi-species genome. arXiv [q-bio.GN] (2023).

18. Dalla-Torre, H., et al. The Nucleotide Transformer: Building and evaluating robust foundation models for human genomics. bioRxiv 2023.01.11.523679 (2023) doi:10.1101/2023.01.11.523679.

19. Bellot, P., de Los Campos, G. & Pérez-Enciso, M. Can deep learning improve genomic prediction of complex human traits? Genetics 210, 809–819 (2018).

20. Badré, A., Zhang, L., Muchero, W., Reynolds, J. C. & Pan, C. Deep neural network improves the estimation of polygenic risk scores for breast cancer. J. Hum. Genet. 66, 359–369 (2021).

21. Zhou, X. et al. Deep learning-based polygenic risk analysis for Alzheimer’s disease prediction. Commun. Med. (Lond.) 3, 49 (2023).

22. van Hilten, A. et al. GenNet framework: interpretable deep learning for predicting phenotypes from genetic data. Communications Biology 4, 1–9 (2021).

23. Li, H., Zeng, J., Snyder, M. P. & Zhang, S. Modeling gene interactions in polygenic prediction via geometric deep learning. Genome Res. 35, 178–187 (2025).

24. Georgantas, C., Kutalik, Z. & Richiardi, J. Deep learning for polygenic risk prediction. medRxiv 2024.04.19.24306079 (2024) doi:10.1101/2024.04.19.24306079.

25. Peng, J. et al. DeepRisk: A deep learning approach for genome-wide assessment of common disease risk. Fundam Res 4, 752–760 (2024).

26. Lee, I. et al. Mechanistic genotype-phenotype translation using hierarchical transformers. bioRxiv 2024.10.23.619940 (2024) doi:10.1101/2024.10.23.619940.

27. Newcombe, P. J., Nelson, C. P., Samani, N. J. & Dudbridge, F. A flexible and parallelizable approach to genome-wide polygenic risk scores. Genet. Epidemiol. 43, 730–741 (2019).

28. Lloyd-Jones, L. R. et al. Improved polygenic prediction by Bayesian multiple regression on summary statistics. Nat. Commun. 10, 5086 (2019).

29. Ge, T., Chen, C.-Y., Ni, Y., Feng, Y.-C. A. & Smoller, J. W. Polygenic prediction via Bayesian regression and continuous shrinkage priors. Nat. Commun. 10, 1776 (2019).

30. Privé, F., Arbel, J. & Vilhjálmsson, B. J. LDpred2: better, faster, stronger. Bioinformatics 36, 5424–5431 (2021).

31. Azodi, C. B., Tang, J. & Shiu, S.-H. Opening the black box: Interpretable machine learning for geneticists. Trends Genet. 36, 442–455 (2020).

32. Watson, D. S. Interpretable machine learning for genomics. Hum. Genet. 141, 1499–1513 (2022).

33. Kuenzi, B. M. et al. Predicting Drug Response and Synergy Using a Deep Learning Model of Human Cancer Cells. Cancer Cell 38, 672–684.e6 (2020).

34. Battaglia, P. W., et al. Relational inductive biases, deep learning, and graph networks. arXiv [cs.LG] (2018).

35. Ewing, C. M. et al. Germline mutations in HOXB13 and prostate-cancer risk. N. Engl. J. Med. 366, 141–149 (2012).

36. Wei, J. et al. Germline HOXB13 G84E mutation carriers and risk to twenty common types of cancer: results from the UK Biobank. Br. J. Cancer 123, 1356–1359 (2020).

37. Kron, K. J. et al. TMPRSS2-ERG fusion co-opts master transcription factors and activates NOTCH signaling in primary prostate cancer. Nat. Genet. 49, 1336–1345 (2017).

38. Wang, J., Cai, Y. & Ittmann, M. TMPRSS2/ERG Fusions. in Encyclopedia of Cancer 2986–2988 (Springer Berlin Heidelberg, Berlin, Heidelberg, 2008).

39. Cancer Genome Atlas Research Network. The molecular taxonomy of primary prostate cancer. Cell 163, 1011–1025 (2015).

40. Abraham, G., Qiu, Y. & Inouye, M. FlashPCA2: principal component analysis of Biobank-scale genotype datasets. Bioinformatics 33, 2776–2778 (2017).

41. Helfand, B. T., Kearns, J., Conran, C. & Xu, J. Clinical validity and utility of genetic risk scores in prostate cancer. Asian J. Androl. 18, 509–514 (2016).

42. Pagadala, M. S. et al. PRState: Incorporating genetic ancestry in prostate cancer risk scores for men of African ancestry. BMC Cancer 22, 1289 (2022).

43. Richard, E. L. et al. Markers of kidney function, genetic variation related to cognitive function, and cognitive performance in the UK Biobank. BMC Nephrol. 23, 159 (2022).

44. Pagadala, M. et al. Germline modifiers of the tumor immune microenvironment implicate drivers of cancer risk and immunotherapy response. Nat. Commun. 14, 2744 (2023).

45. Amos, C. I. et al. The OncoArray Consortium: A Network for Understanding the Genetic Architecture of Common Cancers. Cancer Epidemiol. Biomarkers Prev. 26, 126–135 (2017).

46. McCarroll, S. A. et al. Integrated detection and population-genetic analysis of SNPs and copy number variation. Nat. Genet. 40, 1166–1174 (2008).

47. Bycroft, C. et al. The UK Biobank resource with deep phenotyping and genomic data. Nature 562, 203–209 (2018).

48. Das, S. et al. Next-generation genotype imputation service and methods. Nat. Genet. 48, 1284–1287 (2016).

49. Kuhn, R. M., Haussler, D. & Kent, W. J. The UCSC genome browser and associated tools. Brief. Bioinform. 14, 144–161 (2013).

50. Conti, D. V. et al. Trans-ancestry genome-wide association meta-analysis of prostate cancer identifies new susceptibility loci and informs genetic risk prediction. Nat. Genet. 53, 65–75 (2021).

51. Talwar, J. V., Klie, A., Pagadala, M. S. & Carter, H. GRIEVOUS: your command-line general for resolving cross-dataset genotype inconsistencies. Bioinformatics 40, btae489 (2024).

52. Darcet, T., Oquab, M., Mairal, J. & Bojanowski, P. Vision Transformers Need Registers. arXiv [cs.CV] (2023).

53. Wang, Q., et al. Learning deep Transformer models for machine translation. ACL (2019).

54. Baevski, A. & Auli, M. Adaptive input representations for neural language modeling. ICLR (2019).

55. Shazeer, N. GLU Variants Improve Transformer. arXiv [cs.LG] (2020).

56. Lee, S., Lee, S. & Song, B. C. Improving vision transformers to learn small-size dataset from scratch. IEEE Access 10, 123212–123224 (2022).

57. Loshchilov, I. & Hutter, F. Decoupled weight decay regularization. ICLR (2019).

58. Choi, S. W. & O’Reilly, P. F. PRSice-2: Polygenic Risk Score software for biobank-scale data. Gigascience 8, (2019).

59. Privé, F., Albiñana, C., Arbel, J., Pasaniuc, B. & Vilhjálmsson, B. J. Inferring disease architecture and predictive ability with LDpred2-auto. Am. J. Hum. Genet. 110, 2042–2055 (2023).

60. Akiba, T., Sano, S., Yanase, T., Ohta, T. & Koyama, M. Optuna: A Next-generation Hyperparameter Optimization Framework. in Proceedings of the 25th ACM SIGKDD International Conference on Knowledge Discovery & Data Mining 2623–2631 (Association for Computing Machinery, New York, NY, USA, 2019).

61. Chefer, H., Gur, S. & Wolf, L. Transformer Interpretability Beyond Attention Visualization. in 2021 IEEE/CVF Conference on Computer Vision and Pattern Recognition (CVPR) 782–791 (IEEE, 2021).

62. Abnar, S. & Zuidema, W. Quantifying attention flow in transformers. arXiv [cs.LG] (2020).

63. Chefer, H., Gur, S. & Wolf, L. Generic attention-model explainability for interpreting bi-modal and encoder-decoder transformers. in 2021 IEEE/CVF International Conference on Computer Vision (ICCV) (IEEE, 2021). doi:10.1109/iccv48922.2021.00045.

64. Voita, E., Talbot, D., Moiseev, F., Sennrich, R. & Titov, I. Analyzing multi-head self-attention: Specialized heads do the heavy lifting, the rest can be pruned. Association for Computational Linguistics (2019).

65. 1000 Genomes Project Consortium et al. A global reference for human genetic variation. Nature 526, 68–74 (2015).

66. DeLong, E. R., DeLong, D. M. & Clarke-Pearson, D. L. Comparing the areas under two or more correlated receiver operating characteristic curves: a nonparametric approach. Biometrics 44, 837–845 (1988).

67. Benafif, S., Kote-Jarai, Z., Eeles, R. A. & PRACTICAL Consortium. A review of prostate cancer genome-wide association studies (GWAS). Cancer Epidemiol. Biomarkers Prev. 27, 845–857 (2018).

68. Eeles, R. et al. The genetic epidemiology of prostate cancer and its clinical implications. Nat. Rev. Urol. 11, 18–31 (2014).

69. Thomas, G. et al. Multiple loci identified in a genome-wide association study of prostate cancer. Nat. Genet. 40, 310–315 (2008).

70. Eeles, R. A. et al. Multiple newly identified loci associated with prostate cancer susceptibility. Nat. Genet. 40, 316–321 (2008).

71. Abrahamsson, P. A., Lilja, H., Falkmer, S. & Wadström, L. B. Immunohistochemical distribution of the three predominant secretory proteins in the parenchyma of hyperplastic and neoplastic prostate glands. Prostate 12, 39–46 (1988).

72. McInnes, L., Healy, J. & Melville, J. UMAP: Uniform Manifold Approximation and Projection for Dimension Reduction. arXiv [stat.ML] (2018).

73. Xu, J. et al. HOXB13 is a susceptibility gene for prostate cancer: results from the International Consortium for Prostate Cancer Genetics (ICPCG). Hum. Genet. 132, 5–14 (2013).

74. Adamo, P. & Ladomery, M. R. The oncogene ERG: a key factor in prostate cancer. Oncogene 35, 403–414 (2016).

75. Luedeke, M. et al. Prostate cancer risk regions at 8q24 and 17q24 are differentially associated with somatic TMPRSS2:ERG fusion status. Hum. Mol. Genet. 25, 5490–5499 (2016).

76. Beke, L., Nuytten, M., Van Eynde, A., Beullens, M. & Bollen, M. The gene encoding the prostatic tumor suppressor PSP94 is a target for repression by the Polycomb group protein EZH2. Oncogene 26, 4590–4595 (2007).

77. Chang, B.-L. et al. Fine mapping association study and functional analysis implicate a SNP in MSMB at 10q11 as a causal variant for prostate cancer risk. Hum. Mol. Genet. 18, 1368–1375 (2009).

78. Kote-Jarai, Z. et al. Mutation analysis of the MSMB gene in familial prostate cancer. Br. J. Cancer 102, 414–418 (2010).

79. Cruz, D. F., Farinha, C. M. & Swiatecka-Urban, A. Unraveling the function of lemur Tyrosine Kinase 2 network. Front. Pharmacol. 10, 24 (2019).

80. Shah, K. & Bradbury, N. A. Lemur Tyrosine Kinase 2, a novel target in prostate cancer therapy. Oncotarget 6, 14233–14246 (2015).

81. Geng, C. et al. Prostate cancer-associated mutations in speckle-type POZ protein (SPOP) regulate steroid receptor coactivator 3 protein turnover. Proc. Natl. Acad. Sci. U. S. A. 110, 6997–7002 (2013).

82. Samueli, B. et al. Histopathologic and molecular characterization of IDH-mutant prostatic adenocarcinoma. Mod. Pathol. 38, 100616 (2025).

83. Mucci, L. A. et al. Familial Risk and Heritability of Cancer Among Twins in Nordic Countries. JAMA 315, 68–76 (2016).

84. Hjelmborg, J. B. et al. The heritability of prostate cancer in the Nordic Twin Study of Cancer. Cancer Epidemiol. Biomarkers Prev. 23, 2303–2310 (2014).

