## Supplementary Figures for "VADEr: Vision Transformer-Inspired Framework for Polygenic Risk Reveals Underlying Genetic Heterogeneity in Prostate Cancer"

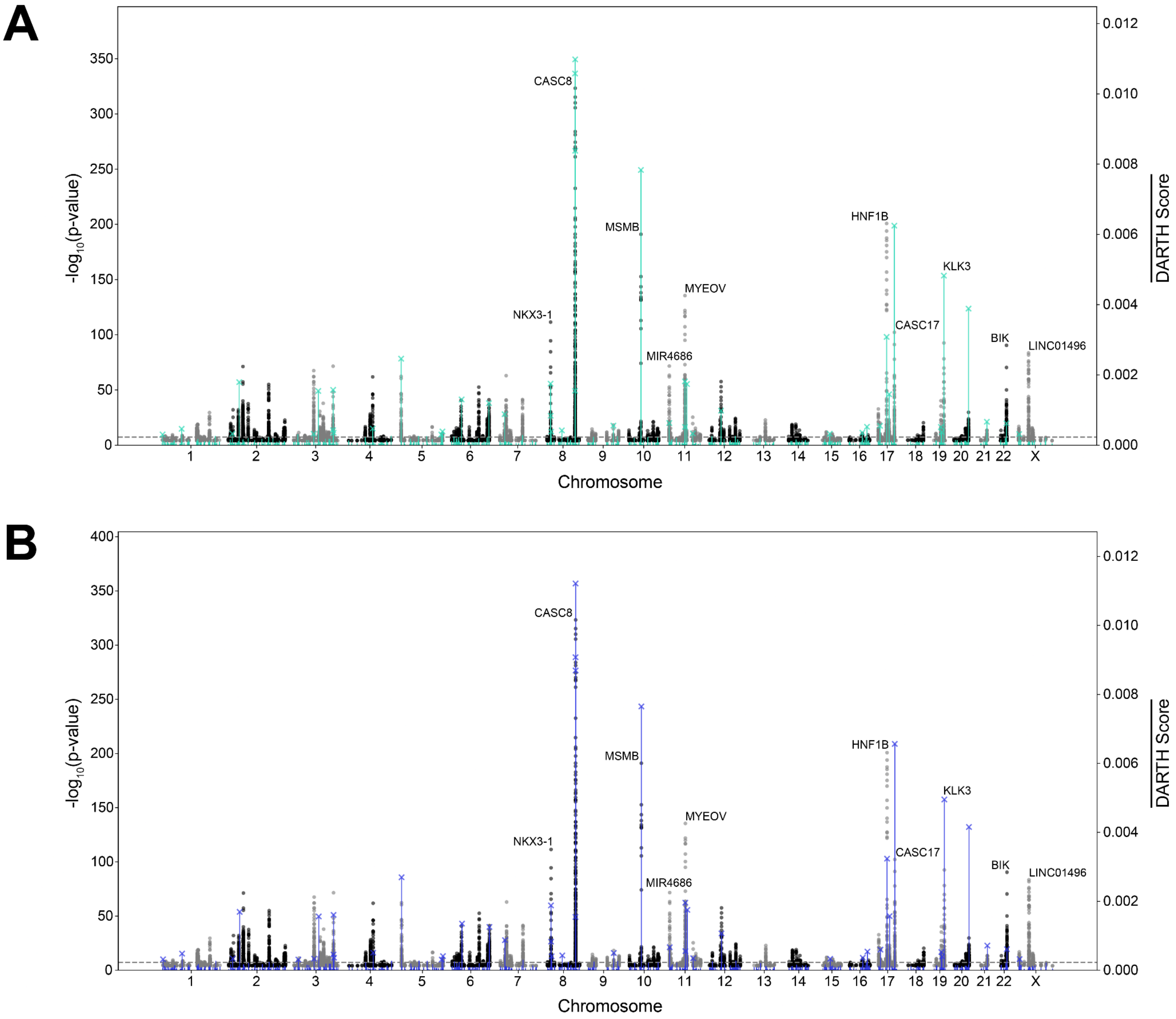


**Supplementary Figure 1:** DARTH Manhattan plots for A) the entire ELLIPSE validation set (n = 18,329) and B) the European ancestry subset of the ELLIPSE validation set (n = 16,513). Colored turquoise and dark blue regions in A) and B) correspond to overlaid per patch average DARTH scores.

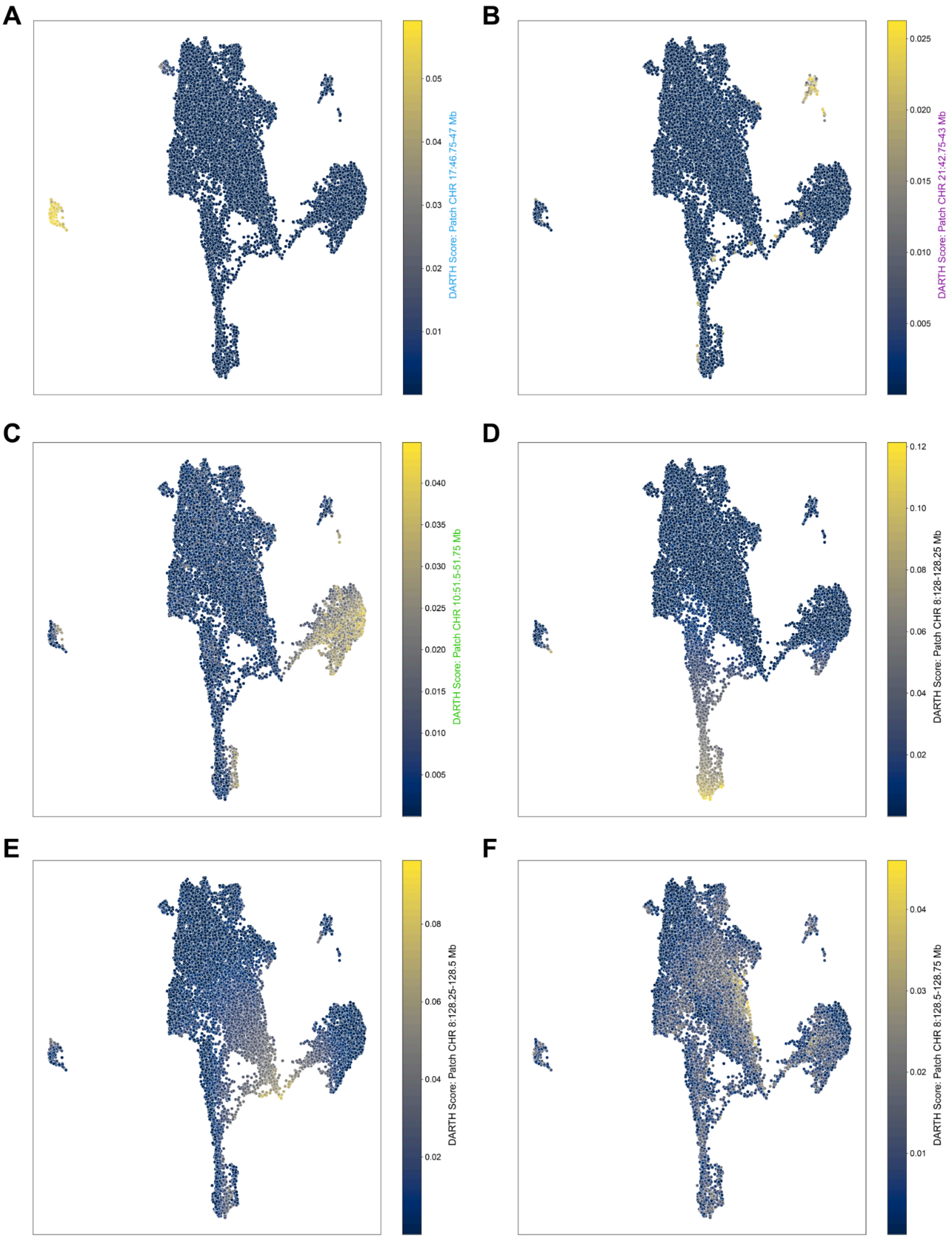
A
DARTH Score: Patch CHR 17:46.75-47 Mb
0.01
0.02
0.03
0.04
0.05
B
DARTH Score: Patch CHR 21:42.75-43 Mb
0.005
0.010
0.015
0.020
0.025
C
DARTH Score: Patch CHR 10:51.5-51.75 Mb
0.005
0.010
0.015
0.020
0.025
0.030
0.035
0.040
D
DARTH Score: Patch CHR 8:128-128.25 Mb
0.02
0.04
0.06
0.08
0.10
0.12
E
DARTH Score: Patch CHR 8:128.25-128.5 Mb
0.02
0.04
0.06
0.08
F
DARTH Score: Patch CHR 8:128.5-128.75 Mb
0.01
0.02
0.03
0.04

**Supplementary Figure 2:** Projecting personalized patch-specific DARTH scores across the UMAP dimensions for A) the top cluster 1 *HOXB13* patch spanning 46.75-47 Mb (GRCh37) on chromosome 17 B) the top cluster 2 *TMPRSS2* patch spanning 42.75-43 Mb on chromosome 21 C) the top cluster 3 *MSMB* patch spanning 51.5-51.75 Mb on chromosome 10 and D-F) the top 3 patches with the highest global average DARTH score which span consecutively from 128-128.75 Mb on chromosome 8.

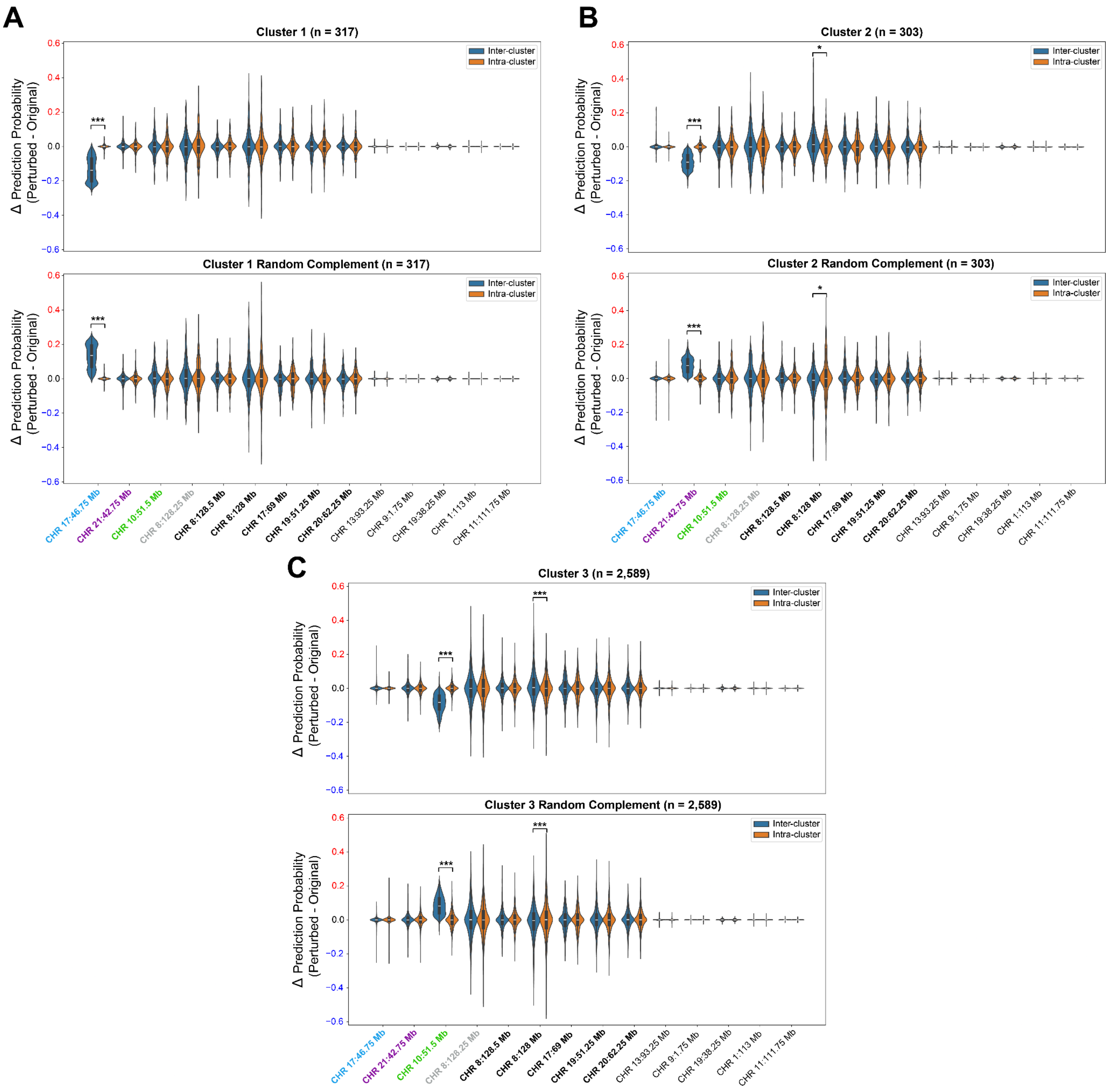
A
Cluster 1 (n = 317)
Cluster 1 Random Complement (n = 317)
B
Cluster 2 (n = 303)
Cluster 2 Random Complement (n = 303)
C
Cluster 3 (n = 2,589)
Cluster 3 Random Complement (n = 2,589)
Inter-cluster
Intra-cluster
Δ Prediction Probability
(Perturbed - Original)
***
*
0.6
0.4
0.2
0.0
−0.2
−0.4
−0.6
CHR 17:46.75 Mb
CHR 21:42.75 Mb
CHR 10:51.5 Mb
CHR 8:128.25 Mb
CHR 8:128.5 Mb
CHR 8:128 Mb
CHR 17:69 Mb
CHR 19:51.25 Mb
CHR 20:62.25 Mb
CHR 13:93.25 Mb
CHR 9:1.75 Mb
CHR 19:38.25 Mb
CHR 1:113 Mb
CHR 11:111.75 Mb

**Supplementary Figure 3:** Quantifying patch contributions to VADEr predicted disease risk through patch genotype shuffling in the UKBB. Patches shown (x-axis) include the top DARTH score patch identified for each cluster (colored accordingly), the top 5 global average DARTH score patches (**bolded**), and the bottom 5 global average DARTH score patches. A) Only genotype shuffling of the identified top cluster 1 patch between populations (compared to within a population) resulted in significant deviations in risk prediction, with a 13.79% decrease (top) and 13.52% increase (bottom) observed across groups. B) Genotype shuffling of the identified top cluster 2 patch between populations resulted in significant decreases of 8.97% (top) and increases of 7.51% (bottom) in VADEr prediction probability. The CASC19 containing patch beginning at chromosome 8:128 Mb also showed modest but significant differences between shuffling methodologies in each group ($p_{cluster2}$ = 0.014; $p_{complement}$ = 0.018). C) Genotype shuffling of the identified top cluster 3 patch between populations resulted in significant decreases of 8.35% (top) and increases of 8.10% (bottom) in VADEr prediction probability. Similar to cluster 2, the patch beginning at chromosome 8:128 Mb also showed modest but significant differences between shuffling methodologies in each population ($p_{cluster3} = 1.7 \times 10^{-4}$; $p_{complement} = 1.5 \times 10^{-4}$).

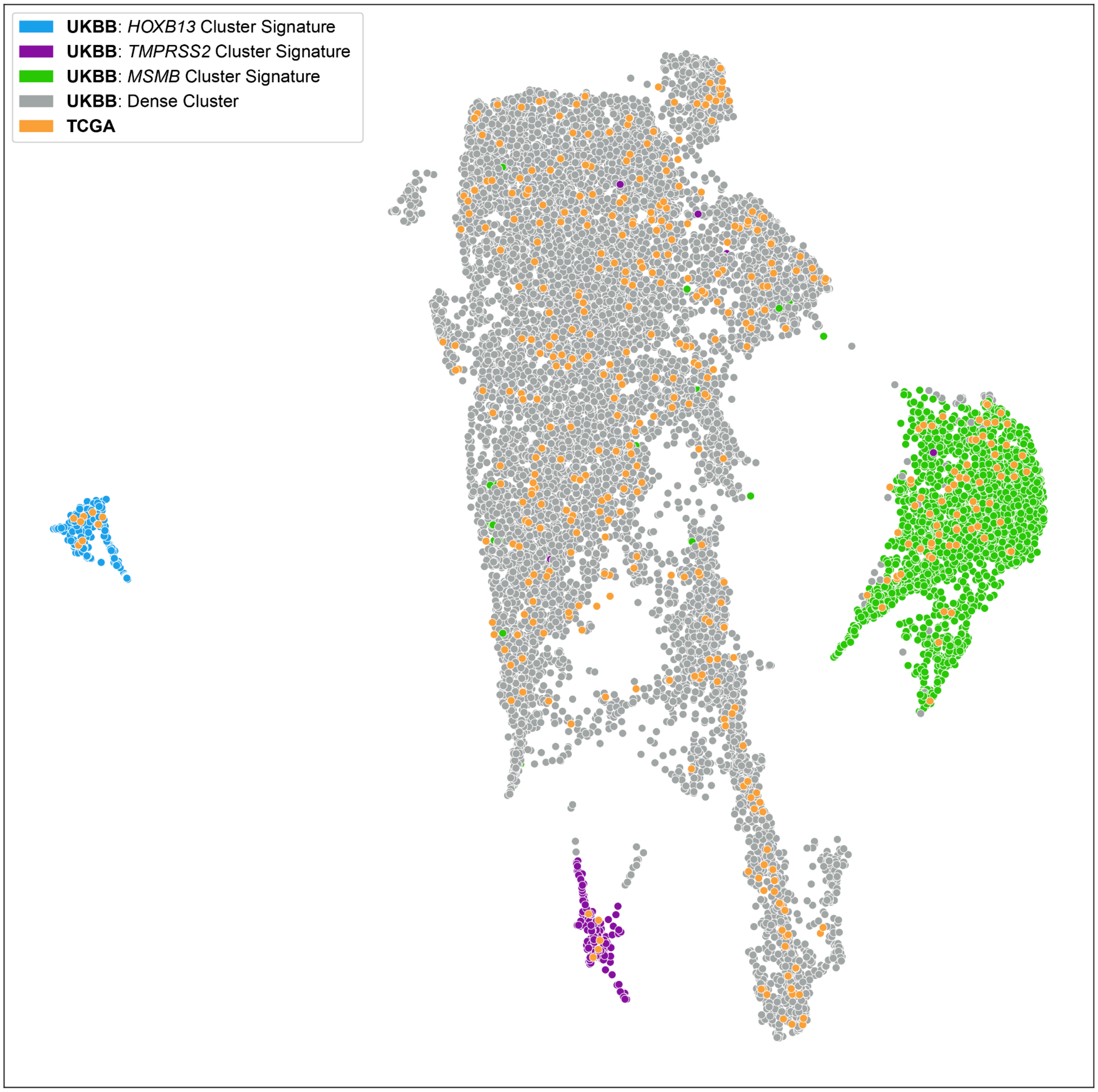


**Supplementary Figure 4:** Joint UMAP projection of DARTH scores in TCGA and UKBB individuals. UKBB individuals are colored according to their identified cluster signature.
